# Stress vulnerability and resilience in children facing COVID-19-related discrimination: A quasi-experimental study using polygenic, brain, and sociodemographic data

**DOI:** 10.1101/2024.03.04.24303643

**Authors:** Jinwoo Yi, Eunji Lee, Bo-Gyeom Kim, Gakyung Kim, Yoonjung Yoonie Joo, Jiook Cha

## Abstract

During the pandemic, perceived COVID-19-related discrimination aggravated children’s stress levels. The remaining question is to evaluate the individual variability in these effects and to identify vulnerable or resilient populations and why. Using the Adolescent Brain and Cognitive Development dataset (*N* = 1,116) and causal machine learning approach – Generalized Random Forest, we examined the average and individual treatment effects of perceived discrimination on stress levels immediately and six months later. Their variability and key factors were also assessed. We observed significant variability in the acute effects of perceived discrimination across children and pinpointed the frontotemporal cortical volume and white matter connectivity (streamline counts) as key factors of stress resilience and vulnerability. The variability of these neurostructural factors partially originated from the environmental and genetic attributes. The finding was replicated in held-out samples (*N* = 2,503). Our study has the potential for personalized prescriptive modeling to prevent children’s future psychopathology after the pandemic.

## Introduction

Despite the World Health Organization marking the end of COVID-19 as a global health emergency^1^, the mental health impacts stemming from the pandemic-induced social injustices are likely to persist^2–5^. Though exposure to moderate stress during developmental stages can enhance an individual’s behavioral and neurobiological resilience^6–8^, excessive or chronic stress during these critical periods can lead to neuronal changes and result in long-term psychopathological condition^9–11^. Consequently, there is a concern that children who experienced COVID-19-related discrimination may face a higher risk of developing mental health disorders, contingent upon the intensity and duration of the distress caused. This concern warrants a thorough investigation to determine the acute and sustained stress impacts of such discrimination and identify which children are particularly susceptible or resilient to these stressors. The urgency for this research is highlighted by the unique stressors presented by the pandemic, such as school closures^12^ and quarantine measures^13^, alongside reports from children across various countries perceiving discrimination related to COVID-19, including racism and “COVID-shame”^14–17^.

In our study, we utilize a comprehensive approach that integrates genetic, sociodemographic, and neuroanatomical data to explore both the acute and sustained effects of perceived COVID-19-related discrimination on stress levels, aiming to pinpoint factors associated with either vulnerability or resilience. Previous studies in animal models have highlighted the neurobiological mechanisms underlying stress vulnerability and resilience, underscoring the importance of genetic and environmental interactions in influencing neurostructural diversity^18–22^. However, human research in this area has often been limited by the absence of longitudinal, multimodal data and sophisticated analytical frameworks, typically focusing on the statistical significance of individual stress moderators. This traditional method has identified numerous predictors of stress response across different contexts, such as socioeconomic status^23, 24^, social support^25, 26^, structural characteristics of the prefrontal cortex^27–29^, and polygenic risk scores for psychiatric conditions^30^. Yet, accurately modeling an individual’s stress response, considering the complex interplay of multiple factors, and identifying the main contributors to resilience or vulnerability remain challenging. This complexity has also led to inconsistent findings, such as the debated protective role of community support against social stressors in adolescents^31–33^ and the contested significance of amygdala morphology^29, 34–36^ and polygenic risk scores^37^. To address these challenges, our research aims to analyze the multivariate interactions among diverse factors, utilizing the extensive Adolescent Brain and Cognitive Development (ABCD) dataset, the largest longitudinal study of children in the United States^38^.

To efficiently and rigorously handle this extensive dataset, we employed the Generalized Random Forest (GRF) method^39^, a cutting-edge causal machine learning technique. GRF excels in estimating both average and individual treatment effects of a treatment variable (in this case, perceived discrimination) on an outcome variable (stress levels) considering various covariates. By utilizing potential outcome frameworks^40^, GRF enables us to explore counterfactual scenarios (e.g., ‘if a child who did not perceive the discrimination experienced it, how much will that child’s treatment effect be?’ and vice versa), offering a more nuanced insight into the impacts of perceived discrimination than traditional statistical methods. This approach is particularly adept at assessing variability in treatment effects among individuals, as it uses the random forest’s greedy algorithm to optimize tree splits, thereby highlighting complex interactions among covariates and identifying key factors related to stress vulnerability (higher effects) and resilience (lower effects). We expect that this quasi-experimental method, supported by a multimodal dataset, will not only contribute to our understanding of human stress responses during the pandemic but also help identify populations vulnerable to perceived COVID-19-related discrimination, laying the groundwork for personalized interventions.

This study addresses three main questions: First, we aim to estimate the acute and sustained average treatment effects of perceived COVID-19-related discrimination on children’s stress levels. Second, we seek to explore individual differences in the acute and sustained treatment effects of perceived discrimination. Lastly, we intend to pinpoint the principal factors of resilience and vulnerability, focusing on individual treatment effects that demonstrate significant variability among children.

## Results

We examined the acute and sustained effects of perceived discrimination on 1,116 children (**Fig 1a** and **Table 1**) using GRF. We aimed to assess how perceived COVID-19- related discrimination (‘treatment’) influenced children’s stress levels, both immediately after the perception of discrimination (‘acute outcome’) and approximately six months later (‘sustained outcome’). To account for individual variability in these effects while considering the interactions among diverse modalities and controlling the confounders between the treatment and outcome variables as much as possible, we used a comprehensive set of 250 covariates derived from an extensive literature review (see ‘Methods’). These covariates encompassed multimodal data, including polygenic scores of psychiatric diseases, environmental, neuroanatomical, psychological, and socioeconomic information (**Supplementary Table 1**). All covariates were measured at least four months before the discrimination report (**Fig 1b**; see ‘Methods’). To mitigate any confounding influences due to site-specific variations in the covariates, outcomes, and treatment variables, we used the data acquisition site as a clustering variable of GRF following the ref^41^.

**Fig 1.**
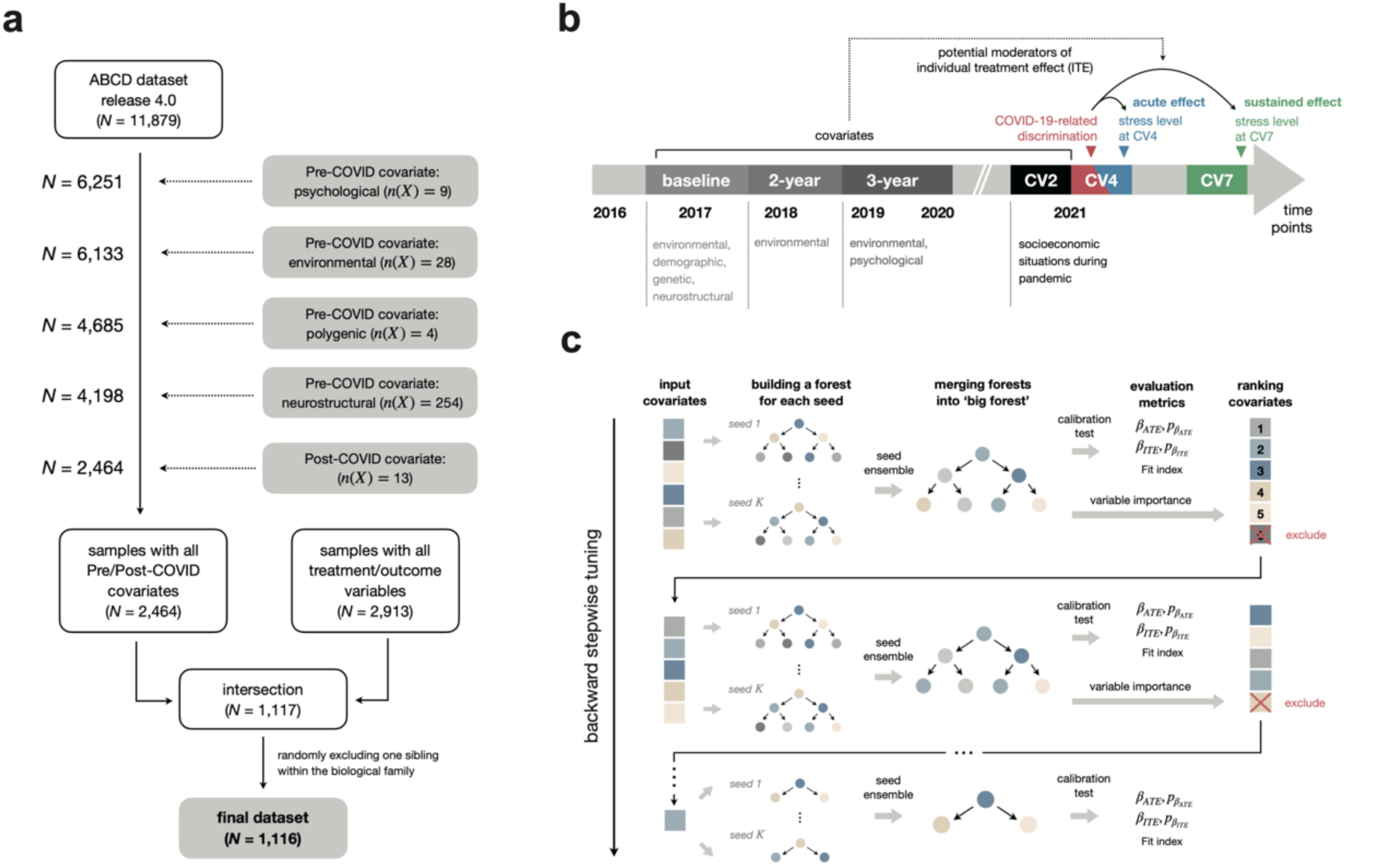
Schematic frameworks for samplings, data collection, and GRF model fitting. **a,** The sampling procedures. **b,** The measurement time points of each variable domain used in the GRF analysis. ‘CV’ is the abbreviation of data acquisition time point during the COVID-19 pandemic. The perceived discrimination reported at CV4 (11/2020 ∼ 02/2021) timepoint was used as a treatment variable. To estimate acute and sustained treatment effects, we defined the perceived stress ratings measured at CV4 and CV7 (05/2021 ∼ 07/2021) time points as acute and sustained outcome variables, respectively. Covariates include 250 variables collected at the baseline (09/2016 ∼ 09/2017), 2-year follow-up (09/2017 ∼ 11/2018), 3-year follow-up (02/2018 ∼ 02/2020), and CV2 (06/2020 ∼ 11/2020) timepoint. **(C)** The overall procedures of GRF fitting and optimization via backward stepwise tuning.

**Table 1.** Stress levels and demographic characteristics of the samples.

| Variables | Overall | Perceiving COVID-19-related discrimination? |  |
| --- | --- | --- | --- |
|  |  | Yes | No |
| *N* | 1,116 | 86 | 1,030 |
| **Baseline and Pandemic stress levels** |  |  |  |
| Pandemic-related stress levels at the CV4 timepoint (11/2020~02/2021), mean (SD) | 2.125 (1.1) | 2.663 (1.1) | 2.080 (1.1) |
| Pandemic-related stress levels at the CV7 timepoint (05/2021~07/2021), mean (SD) | 1.718 (0.9) | 2.070 (1.1) | 1.688 (0.9) |
| Child Behavior Check List Stress2007 scale *t*-score (02/2018~02/2020), mean (SD) | 52.818 (5.4) | 52.919 (5.3) | 52.810 (5.4) |
| **Baseline sociodemographic characteristics** |  |  |  |
| Age month, mean (SD) | 119.256 (7.4) | 119.930 (7.3) | 119.200 (7.4) |
| Sex, n (%) |  |  |  |
| *Male* | 556 (49.8%) | 37 (43.0%) | 519 (50.4%) |
| *Female* | 560 (50.2%) | 49 (57.0%) | 511 (49.6%) |
| Race, n (%) |  |  |  |
| *White* | 787 (70.5%) | 46 (53.5%) | 741 (71.9%) |
| *Black* | 72 (6.5%) | 9 (10.5%) | 63 (6.1%) |
| *Hispanic* | 144 (12.9%) | 13 (15.1%) | 131 (12.7%) |
| *Asians* | 6 (0.5%) | 1 (1.2%) | 5 (0.5%) |
| *Others* | 107 (9.6%) | 17 (19.8%) | 90 (8.7%) |
| Foreign born, n (%) |  |  |  |
| Yes | 334 (29.9%) | 23 (26.7%) | 311 (30.2%) |
| No | 782 (70.1%) | 63 (73.3%) | 719 (69.8%) |
| Parental income, n (%) |  |  |  |
| ~ \$5,000 | 13 (1.2%) | 2 (2.3%) | 11 (1.1%) |
| \$5,000 ~ \$11,999 | 18 (1.6%) | 1 (1.2%) | 17 (1.7%) |
| \$12,000 ~ \$15,999 | 13 (1.2%) | 1 (1.2%) | 12 (1.2%) |
| \$16,000 ~ \$24,999 | 30 (2.7%) | 4 (4.7%) | 26 (2.5%) |
| \$25,000 ~ \$34,999 | 34 (3.0%) | 5 (5.8%) | 29 (2.8%) |
| \$35,000 ~ \$49,999 | 62 (5.6%) | 8 (9.3%) | 54 (5.2%) |
| \$50,000 ~ \$74,999 | 141 (12.6%) | 13 (15.1%) | 128 (12.4%) |
| \$75,000 ~ \$99,999 | 245 (22.0%) | 20 (23.3%) | 225 (21.8%) |
| \$100,000 ~ \$199,999 | 414 (37.1%) | 24 (27.9%) | 390 (37.9%) |
| \$200,000 ~ | 146 (13.1%) | 8 (9.3%) | 138 (13.4%) |
| **Pandemic sociodemographic characteristics** |  |  |  |
| Wage loss since the pandemic, n (%) |  |  |  |
| Yes | 511 (45.8%) | 48 (55.8%) | 463 (45.0%) |
| No | 605 (54.2%) | 38 (44.2%) | 567 (55.0%) |
| County-level cumulative COVID-19 cases, *K*, mean (SD) | 10529.580 (20273.8) | 23807.350 (23689.0) | 10339.400 (19963.8) |
| County-level cumulative COVID-19 deaths, *K*, mean (SD) | 489.910 (869.9) | 557.6 (916.7) | 484.3 (866.1) |
| County-level unemployment rate, %, mean (SD) | 10.362 (3.6) | 10.392 (4.1) | 10.360 (3.6) |

To optimize the model performance of GRF (i.e., how precisely the model estimates average and individual treatment effects), we searched for the best combination of covariates via ‘backward stepwise tuning’ (**Fig 1c**; see ‘Methods’). This feature selection is important to enhance the model performance since the GRF model with a high ratio of covariates to sample size yields an underestimation of average and individual treatment effects^42^. The iterative approach of backward stepwise tuning produced 250 models each for acute and sustained effect analysis. To determine the ‘best model’ for each analysis, we applied two criteria: (1) passing the ‘calibration test’^42^ and (2) displaying the lowest ‘fit index’. Firstly, the models’ performance was evaluated through a calibration test, assessing the explanatory power of estimated average and individual treatment effects on the outcome variable (see equation (5) in ‘Methods’). The calibration test estimates the coefficient of average (′*β_ITE_*′) and individual treatment effect (′*β_ATE_*′) in the regression model to predict the outcome variable. A model that demonstrates significant coefficients for both measures is considered to have passed the calibration test, indicating precise estimation capabilities. Among the models that passed, those with the lowest fit index were selected as the ’best model’ for their respective analyses, according to our established methodology (see equation (6) in ‘Methods’).

### Perceived COVID-19-related discrimination aggravates children’s stress levels immediately and over time

We tested whether the perceived COVID-19-related discrimination has a significant and measurable impact on average stress levels among children. We estimated the acute and sustained average treatment effects using the GRF model (detailed in ’Methods’ section equation (4)) and conducted a calibration test to gauge the accuracy of our average treatment effect estimates (*β_ATE_*)

Our findings indicate a consistent pattern across models, revealing both acute and sustained increases in stress levels as a result of perceived discrimination (**Fig 2a** and **2b**). To synthesize these results, we performed meta-analyses for both the acute and the sustained models (see equation (7) in ‘Methods’). The combined data show substantial effect sizes with significant results (acute: average treatment effect = .536, 95% CI = [.524, .552], *P* < 2×10^-16^; sustained: average treatment effect = .386, 95% CI = [.368, 403], *P* < 2×10^-16^).

**Fig 2.**
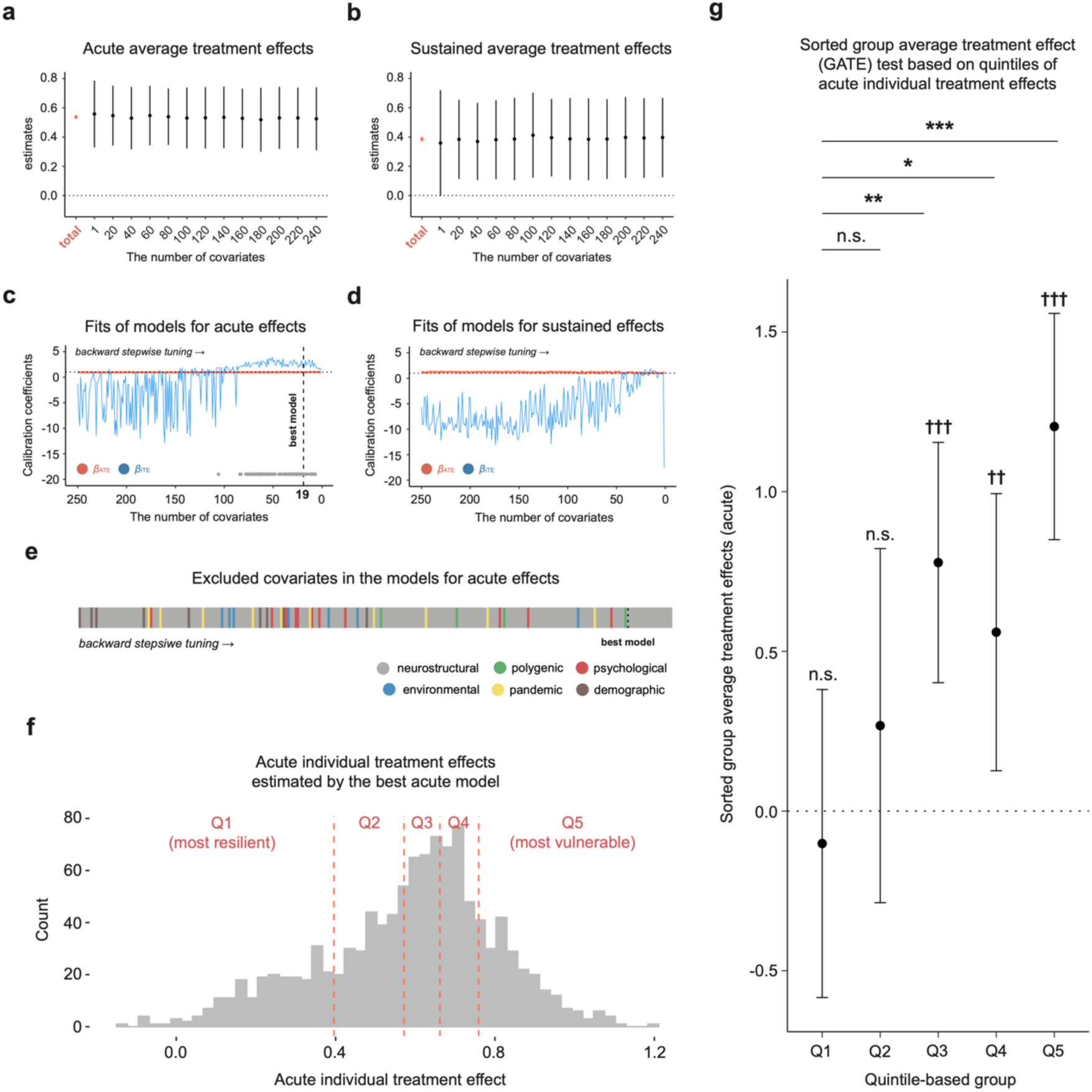
Estimated average treatment effects and assessment of model fit and individual variability. **a,** Estimated acute average treatment effects across models. Error bars show a 95% confidence interval o of estimates. The red bar plot indicates aggregated estimates and confidence intervals across models. From 250 models, only 13 models are visualized here for legibility. **b,** Estimated sustained average treatment effects across models. **c,** Model fit of models for acute effect analysis. The horizontal dashed line indicates the gold standard of the calibration test (i.e., 1). A vertical dashed line means the number of covariates in the best model. Gray points in the bottom right indicate the model showing significant *β_ATE_* and *β_ITE_*. **d,** Model fit of models for sustained effect analysis. **e,** The order of excluded covariates during backward stepwise tuning of models for acute effect analysis. **f,** Distribution of acute individual treatment effects estimated by the best acute model. **g,** Results of quintile-based evaluation of individual variability in acute effects. The error bar denotes a 95% confidence interval of the group-level average treatment effect. Cross symbols above each error bar show the significance of group-level average treatment effects (☨*P* < .05; ☨☨*P* < .01; ☨☨☨*P* < .001). Asterisks above the bar plot indicate the significance of differences in group-level average treatment effects (\**P*_FDR_ < .05; \*\**P*_FDR_ < .01; \*\*\**P*_FDR_ < .001).

All the models for acute and sustained effect analysis consistently displayed point estimates of *β_ATE_* close to the ideal standard of calibration test (i.e., 1), indicating a precise estimation of the average treatment effect (**Fig 2c** and **2d**). Additionally, every model showed a significant *β_ATE_* value at *P*_FDR_ < .05 (**Supplementary Fig 1a**).

These results corroborate the detrimental influence of perceived COVID-19-related discrimination on children’s stress levels, both short-term and persisting over a longer period.

The consistent estimation of the average treatment effect throughout the backward stepwise tuning process suggests that our method of model refinement successfully pinpointed the essential covariates for an accurate average treatment effect estimation.

### Acute effects of perceived discrimination displayed significant individual variability in children, but sustained effects did not

We examined whether individual treatment effects of perceived COVID-19-related discrimination exhibited variability among children. Firstly, we investigated which models showed a significant *β_ITE_* value in the calibration test because the significant *β_ITE_* value indicates not only a modest estimation of individual treatment effects but also the identification of heterogeneity in individual treatment effects^42^. Out of the models analyzed for acute effects, 58 showed a significant *β_ITE_* value (*P*_FDR_ < .05), suggesting detectable individual variability in the acute effects (**Supplementary Fig 1b**). In contrast, none of the sustained effect models reached significance threshold, which may point to either a lack of sufficient covariate information to capture heterogeneity or inherently homogenous sustained effects (**Supplementary Fig 1b**).

Consequently, further evaluation focused solely on acute individual treatment effects. The ’best acute model’ was selected from those that passed the calibration test, identified by 19 neurostructural covariates (**Fig 2e**), yielding a *β_ATE_* of .973 (95% CI = [.614, 1.331], *P*_FDR_ = 3×10^-7^) and a *β_ITE_* of 1.755 (95% CI = [1.007, 2.503], *P*_FDR_ = 2×10^-5^). This model’s robustness was confirmed through a sensitivity analysis using different random seed sets, consistently showing significant *β_ATE_* and *β_ITE_* (**Supplementary Fig 2**). Using this model, we estimated acute individual treatment effects for each subject, categorizing them into quintiles (**Fig 2f**; see ‘Methods’).

We evaluated the individual variability of acute effects by estimating group-level average treatment effects and testing whether the average treatment effects of the other four groups were higher than the estimates of the first group (‘Q1’, with the lowest individual treatment effects) following the ref^42–44^ – called ‘Sorted Group Average Treatment Effect (GATE)’ test. This test revealed a stark contrast in group-level average treatment effects, with the last three quintiles (Q3, Q4, Q5) showing significant acute effects, while the first two quintiles (Q1, Q2) did not (**Fig 2g** and **Supplementary Table 2**). The results demonstrated that except for Q2, all other groups had significantly higher acute average treatment effects compared to Q1 (**Fig 2g** and **Supplementary Table 2**). The consistency of this pattern was also confirmed in GATE tests with different group sizes (**Supplementary Fig 3**). These findings indicate that the ’best acute model’ successfully captured significant individual variability in children’s immediate stress response to perceived discrimination.

### Variability of the frontotemporal morphology is associated with individual differences in acute effects of perceived discrimination

To identify key factors related to individual differences in acute personalized effect of perceived discrimination, we conducted triangulated analyses of covariates from the ‘best acute model’ (**Fig 3a**; see ‘Methods’). The ‘best acute model’ incorporated 19 neurostructural covariates, including four gray matter volume features and 15 white matter streamline count variables in frontotemporal areas.

**Fig 3.**
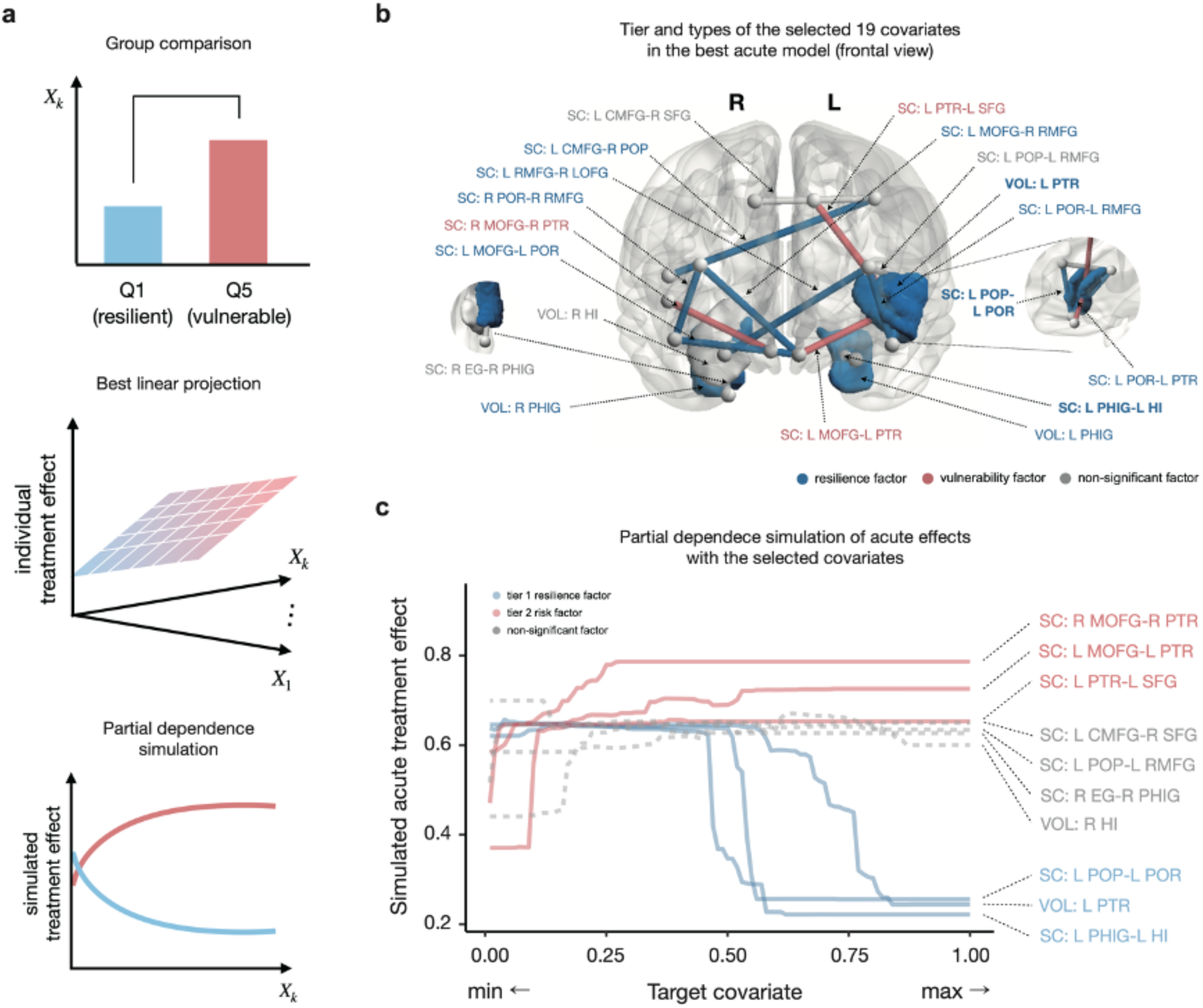
Framework and results of covariate analyses. **a,** The overall procedures of the triangulated covariate analyses. **b,** The tier and types of the selected 19 covariates in the best acute model. The colored region denotes the cortical volume variable, and an edge between two nodes depicts the streamline count variable. The bolded name of covariates indicates the tier 1 factor. **c,** Results of partial dependence simulation. 0 and 1 in the x-axis mean each covariate’s minimum and maximum value, respectively. The y-axis shows the acute treatment effect given each covariate, simulated by the best acute model. Among 12 resilience factors, we only visualized the results of tier 1 factors for legibility. Abbreviation: CMFG: caudal middle frontal gyrus, EG: entorhinal gyrus, HI: hippocampus, LOFG: lateral orbitofrontal gyrus, MOFG: medial orbitofrontal gyrus, PHIG: parahippocampal gyrus, POP: pars opercularis, POR: pars orbitalis, PTR: pars triangularis, RMFG: rostral middle frontal gyrus, SFG: superior frontal gyrus. SC: streamline counts. VOL: cortical volume.

Firstly, we tested statistical differences in each covariate between the most resilient (Q1) and vulnerable (Q5) groups (‘group comparison’). We identified 12 resilience factors (i.e., larger in Q1), three vulnerability factors (i.e., larger in Q5), and four non-significant factors. Next, we conducted multiple regression analysis to examine whether each covariate significantly explains the variance of acute individual treatment effects while other features are controlled (‘best linear projection’). This analysis singled out three of the resilience-associated covariates as significant predictors of acute effects (**Supplementary Table 3**).

We then classified covariates in terms of two criteria: (1) the direction of the effects (‘type,’ resilience or vulnerability factors) and (2) the number of significant results in both analyses (‘tier,’ two or one). For instance, the cortical volume of the right parahippocampal gyrus was assigned to the tier 2 resilience factor since it was greater in Q1 than in Q5 but did not show a significant negative coefficient in the best linear projection analysis. This analysis demonstrated three tier 1 resilience factors – the cortical volume of the left pars triangularis, the streamline counts between the left pars opercularis and pars orbitalis, and the streamline counts between the left parahippocampal gyrus and the left hippocampus. Also, nine tier 2 resilience factors, three tier 2 vulnerability factors, and four non-significant ones were identified (**Fig 3b** and **Supplementary Table 3**).

To further probe the individual treatment effects and their dependency on the covariates, we conducted a partial dependence simulation. This was performed by holding the remaining 18 variables at their median values, allowing us to examine each covariate’s impact without assuming linearity or struggling with variable interactions present in group comparisons. This analysis demonstrated that resilience and vulnerability factors are associated with decreases and increases in acute responses, respectively, unlike non-significant factors (**Fig 3c**).

Our comprehensive three-step covariate analysis confirms that the structural attributes of the frontotemporal regions are pivotal in the variability of individual acute stress reactions to perceived discrimination.

### Individual difference in acute effects of perceived discrimination is detected by the frontotemporal structure in the held-out dataset

We assessed the reproducibility of our findings with the held-out samples (*N* = 2,503; see ‘Methods’). The following two questions were tested: (1) Can individual differences in acute responses to perceived discrimination be elucidated by the identified 19 frontotemporal morphology covariates in the held-out samples? (2) Is there congruence between the tier and type classifications of each covariate in the held-out dataset compared to the original?

Firstly, we observed that subsets of frontotemporal variables selected in the original dataset successfully detected variability in acute stress response. Two models from 19 GRF models built in backward stepwise tuning passed the calibration test (i.e., showing significant *β_ATE_* and *β_ITE_* at *P*_FDR_ < .05) (**Supplementary Fig 4a**). Specifically, significant *β_ITE_* indicates that these models captured heterogeneity in acute individual treatment effects. To further assess the individual difference in acute effects further, we performed the GATE test as we did with the original dataset (**Supplementary Fig 4b**). Except for Q4, every group showed significantly larger average treatment effects than Q1 (**Supplementary Fig 4c**). Secondly, the correspondence of tiers and types among the neurostructural variables between the datasets was modestly aligned (tiers: Cramer’s *V* = .344; types: Cramer’s *V* = .415) (**Supplementary Fig 4d**). Overall, the results from the reproducibility evaluation with the held-out dataset support the robustness and generalizability of our findings.

### Key neurostructural factors of individual variability in acute effects of perceived discrimination accommodate environmental and genetic attributes

Our findings revealed the relationship between the frontotemporal structures and acute stress responses to perceived COVID-19-related discrimination. Then, how was this variability of the brain structure shaped? We tested whether the frontotemporal morphometry included genetic and environmental influences. For this, we employed sparse generalized canonical correlation analysis (sgCCA) allowing simultaneously analyze and maximize correlations across multiple variable sets^45^.

We applied sgCCA to the following three sets of variables: 19 neurostructural (selected in the ‘best acute model’), four polygenic risk scores (depression, posttraumatic stress disorder, schizophrenia, and body mass index) and nine environmental variables (parental monitoring, parents’ caregiving family conflicts, child’s prosocial behavior, school engagement, school disengagement, school environment, neighborhood safety, and traumatic episode). To derive directional interpretations from the sgCCA model, we chose nine environmental variables measured at the baseline timepoint among 28 environmental covariates in the main GRF analysis as potential sources of variability in the neurostructural structures. For a robust evaluation, we divided the dataset evenly into discovery and replication sets, maintaining equal proportions of resilience/vulnerability groups (Q1-Q5) through stratification. Using the discovery set, we explored the optimal sparsity hyperparameters and assessed the relationships among blocks with the replication set. Lastly, we tested whether the neurostructural canonical variate modeled in the sgCCA displays significant relationship with the acute individual effects estimated by the best acute model (**Fig 4a**; see ‘Methods’).

**Fig 4.**
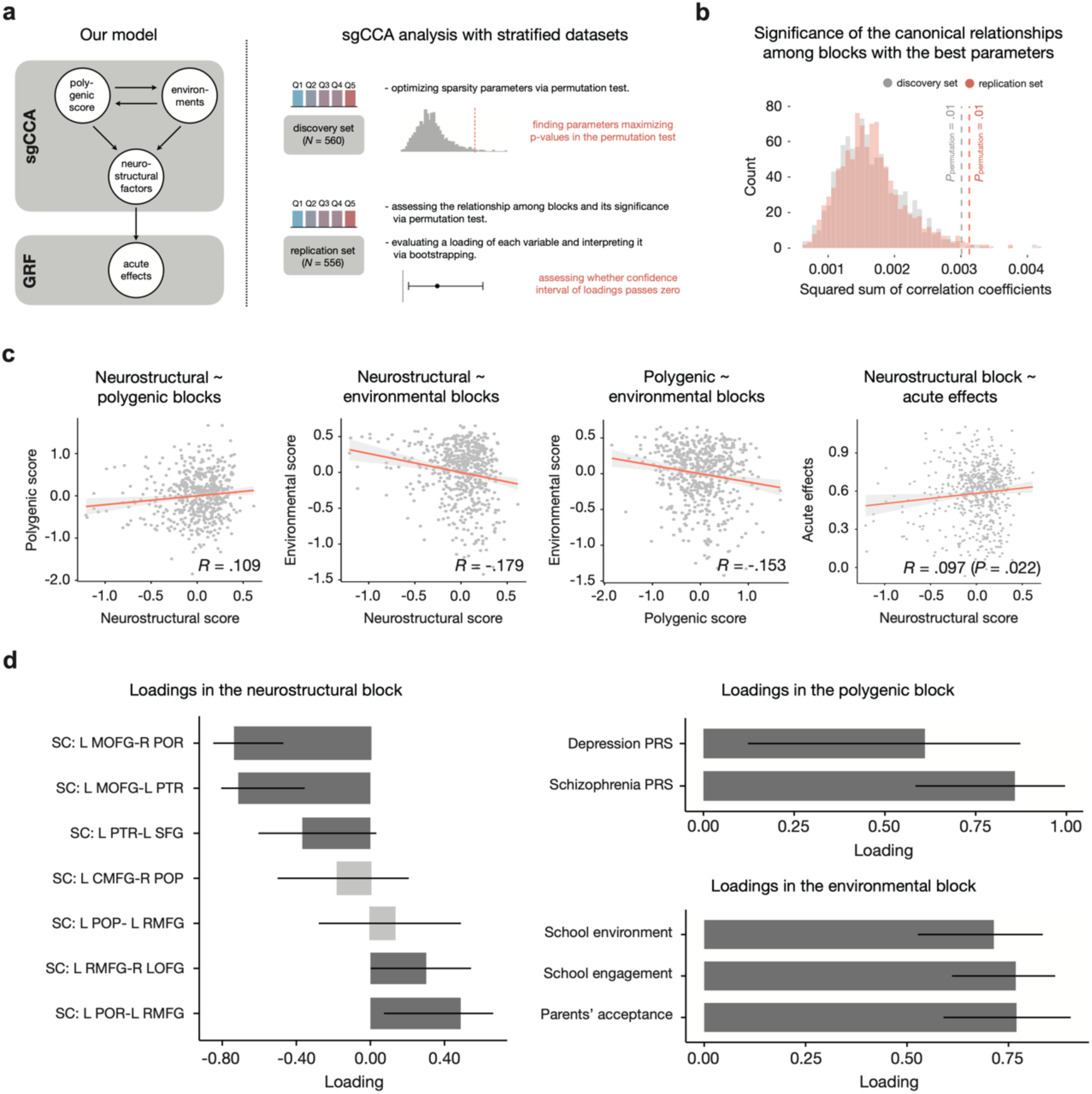
Framework and results of sgCCA modeling with key frontotemporal factors and environmental/polygenic covariates. **a,** Model tested by sgCCA modeling and overall framework. **b,** The significance of the multivariate relationship among blocks in the discovery and replication sets. Statistical significance was assessed via a 1,000 times permutation test. Each histogram shows the null distribution obtained from the permuted sgCCA models. **c,** Multivariate relationship among blocks. *R* means the Pearson correlation coefficient between canonical variates. The Pearson correlation test separately tested the association between the neurostructural block and acute effects. **d,** Loadings of covariates in each block. Each bar denotes the standardized loading. Dark gray color bars exhibited significant loadings. The error bar shows the confidence interval of loadings obtained from the 1,000 times bootstrapping.

The sgCCA results showed a significant relationship among three blocks in the discovery and replication sets (*P*_permutation_ = .01) (**Fig 4b**). The canonical variate of the polygenic scores was positively correlated with the neurostructural variate (*r* = .109), whereas the environmental variate exhibited a negative influence (*r* = -.179). Polygenic and environmental variates were negatively correlated (*r* = -.153). Further, the neurostructural canonical variate derived from the sgCCA model showed a significant positive correlation with acute individual treatment effects (*r* = .097, *P* = .022), indicating its predictive relevance (**Fig 4c**).

Significant loadings within each block were identified, indicating key variables (**Fig 4d**). In the neurostructural block, negative loadings were observed for white matter streamline counts between the left medial orbitofrontal gyrus and right pars opercularis, streamline counts between the left medial orbitofrontal gyrus and left pars triangularis, and streamline counts between the left pars triangularis and left superior frontal gyrus. Positive lodaings were shown in the streamline counts between the left rostral middle frontal gyrus and right orbitofrontal gyrus and streamline counts between the left pars orbitalis and left rostral middle frontal gyrus. In the polygenic scores block, positive loadings were found for schizophrenia and depression scores presented. In the environmental block, positive loadings were found for parents’ acceptance and school environment/engagement (see **Supplementary Table 4**).

The sgCCA findings suggest that the variability in frontotemporal structures is rooted in a complex interplay of genetic and environmental factors, and that these characteristics are predictive of acute stress responses to perceived discrimination associated with COVID-19.

## Discussion

We investigated the acute and sustained impacts of perceived COVID-19-related discrimination on children’s stress levels and identified the neuronal structure variables underlying children’s resilience or vulnerability to the stressor. We found that perceived discrimination elevates stress level both immediately (acute effect) and at six-month follow-up (sustained effect). While our model captured the variability in acute stress reactions, it noted no notable differences in sustained stress outcomes. In estimating the acute effect in each individual, 20% of children experienced acute responses twice that of population-level estimates (vulnerable group, Q5). Contrarily, 40% of children showed no significant stress responses to perceived discrimination (the first two resilient groups, Q1 and Q2). The frontotemporal morphological features emerged as critical indicators of acute stress responses, a finding corroborated by a larger held-out dataset. These neurostructural factors were significantly associated with environmental and genetic factors, suggesting that a gene-environment interplay shapes the neurobiological underpinnings of stress resilience and vulnerability.

Our quasi-experimental approach to estimating acute average treatment effects supports that such stressor during the pandemic aggravates stress levels in children, aligning with the previous studies^2, 16, 46^. Also, at the individual level, we modeled all sample’s acute effects considering the complex interplay among diverse domains covariates and captured significant individual variability. The polarized pattern of children and adolescents’ immediate stress response to perceived discrimination has also been consistently observed in the existing literature^31, 47–49^. The main difference between our study and previous findings is that while they defined the variability factors (e.g., self-esteem^48^ or coping strategies^31, 49^) a priori and tested significance of differences in acute stress responses between groups, our approach leveraged data-driven sub-grouping based on causal estimates and explored the variability factors in a bottom-up way. This method provides a more nuanced identification of sub-groups, encompassing the multifaceted interactions among variability sources that were challenging to handle with conventional approaches^43, 50^. Furthermore, our framework overcomes the selection bias (i.e., estimating and assessing the variability in the effects of perceived discrimination only with the samples reporting perceived discrimination) via counterfactual modeling with potential outcome frameworks. This approach enables us to obtain less biased estimates of the effects of perceived discrimination and extend the preventive identification of vulnerable populations to children not experiencing discrimination yet.

While our models indicated a significant sustained average treatment effect on children’s stress levels, we observed no significant variability in this impact. This may be unexpected as stress resilience is known to vary among individuals over time^51, 52^. A potential reason for this could be the omission of post-discrimination covariates from our analysis. Since structural changes in the ventromedial prefrontal cortex^53–55^ and hippocampus^56, 57^ following traumatic events are linked to the modulation of stress responses, the absence of post-discrimination neural measurements in the ABCD dataset might prevent the detection of individual sustained response variations. To address this, future research should incorporate the forthcoming neuroanatomical dataset that spans from the instances of discrimination to the resulting stress levels.

Our results are in line with prior studies that have identified structural features in the prefrontal region, particularly the medial prefrontal cortex, as key indicators of stress resilience. This region modulates stress responses through its connections with subcortical-limbic areas via the hypothalamic-pituitary-adrenal (HPA) axis^58–60^. Increased white matter myelination in this region, possibly reflected in the larger cortical volume, may facilitate the transmission efficiency in this emotion regulation and stress resilience^6^. Additionally, we have identified the morphology of the inferior frontal gyrus’s gray and white matter as tier 1 resilience factors, crucial in differentiating between resilient and vulnerable populations^61, 62^. The role of hippocampal complex connectivity as a tier 1 protective factor is a relatively new finding. Functional activity in these regions has been found to inversely correlate with perceived stress during cognitive tasks^63^. While further research is needed to explore the relationship between white matter structure and stress-related brain activations, our data allude that white matter hyperconnectivity within the hippocampal complex could be significant in stress response regulation. The consistency of these findings across different sample sets also underscores the broad applicability of the link between frontotemporal structures and acute stress responses. Furthermore, the variations in covariates’ types between the original and held-out datasets highlight the individual differences in how neural features are engaged during immediate stress processing, reported in the previous findings^64, 65^.

Our sparse generalized canonical correlation analysis (sgCCA) indicates that brain structures adaptively respond to acute stress by integrating individual environmental factors and genetic predispositions toward psychiatric disorders. In our model, environmental and polygenic canonical variates displayed consistent characteristics as resilience and vulnerability factors, respectively. Parental acceptance^66, 67^ and school engagement^68, 69^ which support adaptive stress-coping strategies in children, exhibited significant positive loadings in our sgCCA model. Contrarily, polygenic risk scores of depression and schizophrenia, predicting one’s stress vulnerability at the behavioral level^30, 70^, were also selected and presented positive loadings in the polygenic block. These dual influences on the neurostructural variables are consistent with the epigenetic model of stress response variability observed in animal studies^18–22^. At the microstructural level within the brain, enriched environments are shown to increase ΔFosB expression, which in turn facilitates glutamate transmission from the prefrontal cortex to limbic regions and enhances hypothalamic-pituitary-adrenal (HPA) axis regulation, thereby boosting behavioral resilience^71^. Conversely, isolation in the postnatal developmental stage disrupts prefrontal myelination^72^ and dopaminergic projection^73^ through altered gene expression, leading to chronic maladaptive behaviors. In line with the literature, our results add that gene-environmental interactions would partly shape the frontotemporal circuit, the primary source of individual variability in acute stress responses.

Notably, individual neurostructural variables showed distinct behaviors in univariate versus multivariate analyses within the same dataset. For example, streamline counts between the left medial orbitofrontal gyrus and both the right pars orbitalis and the left pars triangularis presented as distinct entities in group comparisons yet exhibited collective negative associations in the sgCCA model. This pattern hints that neurostructural variables with seemingly opposing roles in stress response might operate cohesively within a circuit at the multivariate level. This underscores the advantage of multivariate methods, such as the sgCCA, in capturing the interdependencies within neural networks that univariate analyses might miss^74–76^. Univariate approaches often overlook the circuit-level projections of neuroendocrine systems that are integral to stress resilience and vulnerability^18–22^, a gap that multivariate approaches can address. Indeed, human neuroimaging studies have found multivariate methods to be more revealing of the connections between neuroanatomical networks and behavioral resilience compared to the conventional univariate approach^77^. Although the results of our threefold covariate analyses support the specific relationship between each neurostructural variable and acute effects even after when handling the covariance among the brain structures, our multivariate modeling also necessitates the circuit-wide interpretation of the frontotemporal network’s engagement in the variability of stress response to perceived discrimination.

Our study’s conclusions must be drawn with caution due to several limitations. Firstly, the potential violations of causal assumptions warrants consideration. The binary treatment variable for the frequency of perceived COVID-19-related discrimination might compromise the stable unit treatment value assumption (SUTVA), as there is inherent variability in the types, frequencies, and intensities of discrimination that children encounter. Hence, certain degrees of individual differences in acute effects might be related to the diverse nature of the perceived discrimination rather than its personalized effects per se. However, previous meta-analytic work found no significant psychological effect differences attributable to types of discrimination^78^. Also, we did not observe any significant differences in the frequency of perceived discrimination across acquisition sites (**Supplementary Fig 5**). The absence of data on discrimination intensity within the ABCD dataset suggests a need for further investigation into how this intensity correlates with psychological outcomes. Secondly, our backward stepwise modeling might not fully account for non-neural covariates in the variability of acute stress responses. Including GRF, forest-based models assign importance based on covariate frequency in tree splitting, potentially sidelining categorical variables with fewer categories^79, 80^. Consequently, certain factors, such as gender or early-life trauma, might be systematically overlooked during model selection. Despite this, our incorporation of environmental variables into the sgCCA modeling revealed significant associations with frontotemporal factors, underscoring the role of environmental factors in stress resilience and vulnerability that may have been underappreciated in the primary GRF analysis. Lastly, the generalizability of our findings needs validation through an independent dataset. For example, Asians, who have reportedly faced frequent COVID-19- related discrimination^81–83^, represent only 0.5% of our study sample. This racial imbalance within the ABCD dataset could limit the representativeness of our results. The absence of longitudinal, multimodal datasets collected before and after the pandemic across varied demographics prevents us from fully addressing this limitation, highlighting the need for future research with a more representative dataset.

Notwithstanding these limitations, our study has significant implications for neuroscience and social science. Firstly, our multimodal approach allows us to identify key neuronal factors that mediate gene-brain-environment interactions, illustrating the complex interplay in human stress response variability. Secondly, our data-driven approach pinpoints essential factors to target individuals vulnerable to discrimination. Unlike traditional studies that focus on populations already experiencing discrimination, our quasi-experimental analysis identifies those potentially at risk before such experiences occur. This approach paves the way for a personalized model of prescriptive analysis that could anticipate and mitigate the psychopathological impact of perceived discrimination, thereby enhancing the prioritization of mental health services. Most importantly, our research highlights the effects of discrimination on children during the COVID-19 pandemic, underscoring the urgent need for societal and healthcare preparedness in the post-pandemic landscape.

## Methods

### Dataset

#### Study population

We used data from the Adolescent Brain Cognitive Development (ABCD) study release 4.0 dataset (http://abcdstudy.org). Out of the 11,879 children in the ABCD dataset, our final sample comprised 1,116 individuals. Our initial pool included 4,198 participants who had ‘pre-COVID’ covariates – encompassing demographic, psychological, environmental, polygenic, and neural variables measured prior to the pandemic – and 6,337 participants with complete ‘post-COVID’ covariates, specifically socioeconomic variables gathered during the pandemic. We then formed a subset of 2,464 participants at the intersection of these two groups. After coding non-informative responses (e.g., “don’t know” or “prefer not to answer”) as missing values, we considered excluding participants with missing data exceeding 10% of the number of covariates. However, this step led to no exclusions. We further refined the sample by intersecting the remaining 2,464 participants with an additional 2,913 who provided complete information on both the treatment (i.e., perceived COVID-19-related discrimination) and two outcomes (i.e., the perceived stress levels immediately and about six months after the discrimination experience). From 1,117 samples from this intersection, we randomly excluded one sibling within the biological family to ensure independent sampling. Finally, we determined 1,116 children as the final population.

#### Covariates

Our study engaged a comprehensive set of covariates to investigate stress vulnerability and resilience among children in response to perceived COVID-19-related discrimination. We incorporated 314 covariates identified through an extensive literature review. These were grouped based on their temporal relation to the pandemic breakout: 301 ‘pre-COVID’ and 13 ‘post-COVID’ covariates.

The ‘pre-COVID’ set consisted of five domains: six demographic, nine psychological, 28 environmental, four polygenic, and 254 neurostructural variables. Demographic variables contained sex, age, born overseas, race, the first caregiver’s income, and their final degree, measured at the baseline timepoint (09/2016 ∼ 09/2017). Psychological measures covered several symptom scores related to mood disorder, baseline stress levels, and emotion regulation strategies, assessed at the 3-year-follow-up timepoint (02/2018 ∼ 02/2020).

Environmental variables captured familial interactions, prosocial behaviors, neighborhood safety, and children’s traumatic experiences, such as the loss of a loved one, reported at the baseline time point. The environment domain also included the perceived discrimination and racial identity, reported at the 2-year-follow-up (09/2017 ∼ 11/2018) and 3-year-follow-up timepoints, respectively. Genetic predispositions to stress response were quantified through polygenic risk scores for traits associated with the stress responses – depressive symptoms, schizophrenia, post-traumatic stress disorder, and body mass index. These scores were calculated from the multi-ethnic ancestry reference panels. Detailed genotyping, computation, and validation procedures of the polygenic risk scores are demonstrated in **Supplementary Note 1**. Lastly, neuronal covariates encompassed 23 gray matter cortical volume variables and 231 white matter streamline count variables. These variables were derived from 11 regions of interest (ROIs) - the superior frontal gyrus, caudal middle frontal gyrus, rostral middle frontal gyrus, pars opercularis, pars triangularis, pars orbitalis, medial orbitofrontal gyrus, lateral orbitofrontal gyrus, entorhinal cortex, parahippocampal gyrus, and hippocampus. Our selection was informed by a review paper that associated frontotemporal morphologies with stress resilience to traumatic experiences^29, 36^. We utilized bilateral cortical volumes from these regions and the whole-brain cortical volume. For a detailed demonstration of preprocessing and estimation of T1 and T2 images in the ABCD study, see the ref^84^. Additionally, we estimated streamline counts for every possible bilateral ROI pair using diffusion-weighted imaging data. The methodologies for modeling tractographies and counting streamlines are detailed in **Supplementary Note 2**. All neuronal variables were determined based on neuroimaging data collected at the baseline time point.

For the ‘post-COVID’ covariates, we utilized 13 COVID-19 Rapid Response Research Survey variables in the ABCD release 4.0 dataset. They reflect children’s reports about familial relationships and parents’ interviews about their economic situations and their child’s history of COVID-19 infection. Additionally, geocoded data included social distancing metrics (e.g., the average amount of time at home), county-level COVID-19 prevalence (e.g., the number of COVID-19 deaths), and labor statistics (e.g., the unemployment rate). All post-COVID variables were sampled at the ‘COVID 2’ (CV2) timepoint (06/2020 ∼ 11/2020).

From the initial covariate set, we removed those with over 10% missing values, including two variables about children’s racial identity and two features regarding family relationships during the pandemic. We imputed missing data for the remaining 310 covariates using the k-nearest neighbors algorithm (*k* = 33; the odd number closest to the square root of the sample size), as implemented by the VIM package^85^. We further refined our covariate set by eliminating those with near-zero variance using the caret package’s ‘nearZeroVar’ function^86^. This preprocessing led to excluding 62 covariates, yielding a final set of 248. Categorical variables were then transformed into dummy variables, culminating in a dataset comprising 250 covariates for our analysis. See **Supplementary Table 1** for detailed information about the final set of covariates.

#### Treatment

Our treatment variable was defined as the self-reported frequency of experiencing racism or other types of COVID-19-related discrimination within the previous week (“Over the past week, I experienced racism or discrimination in relation to coronavirus”). The response was collected at the CV4 (CV4) timepoint (11/2020 ∼ 02/2021). Children responded using a 5-point Likert scale, ranging from 0 (“Never”) to 4 (“Very Frequently”). To facilitate our Generalized Random Forest (GRF) analysis, we converted these responses into a binary format: 0 (“Never”) and 1 (“At least once”). This transformation was the only available setting because higher thresholds for binarization led to a dummy variable with near-zero variance, limiting our analytical options. Therefore, this initial cutoff was the most viable method for binary coding of the treatment variable in our dataset.

#### Outcome

From the 5-item COVID-Related Worry questionnaire in the ABCD study, we used a stress-related measurement as the outcome variable (“In the past 7 days, including today, how stressful have you found the uncertainty COVID-19 presents to be?”). Although the Perceived Stress Scale^87^ measured during the pandemic in ABCD was also available, we did not use this information because its broader time scale (i.e., measuring one’s stressful cognition in the past month rather than week) may disturb the estimation of the effects of perceived discrimination to the stress levels. Responses reported at the CV4 and CV7 (05/2021 ∼ 07/2021) time points permitted us to estimate children’s immediate stress response (acute effect) and about six months later response (sustained effect) to the perceived discrimination. Both metrics were composed of a 5-point Likert scale (i.e., from 1 = “Very Slightly” to 5 = “Extremely”). All variables, including covariates, treatment, and outcome variables, were standardized with the ‘preProcess’ function in the caret package^86^, except for dummy variables of categorical features.

### Generalized Random Forest analysis

#### Overview of Generalized Random Forest

Generalized Random Forest (GRF) is adept at estimating both the average and individual treatment effect for a given treatment, outcome, and covariates^39^. It is designed to encapsulate the heterogeneity in individual treatment effects, thereby facilitating an in-depth evaluation of disparities in individual treatment effects and the identification of key factors.

During tree construction, GRF randomly selects samples and covariates as the ordinary random forest does. Since we used data acquisition site as a clustering feature, it extracts the same number of samples from every cluster in this step. This technique intends to give the equal weight to every cluster and achieve more generalizable prediction to the unseen data^42^. Inspired by the greedy algorithm of the random forest, it optimizes node splits to accentuate differences in treatment effects between the resulted nodes. Treatment effect of the node *L*, *τ_L_*, is estimated with the following residual-on-residual regression:

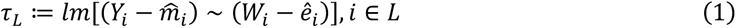

where *i* represents a sample index, *Y* is the outcome variable, *m̂_i_* is the expected outcome given the covariate, *W* is the treatment variable, *ê_i_* is the predicted treatment given the covariate (i.e., propensity score). To procure out-of-sample predictions for *m* and *e* of the sample *i*, GRF establishes ‘regression forest’ separately before fitting the main causal forest model.

GRF posits that samples within the same terminal node share a homogeneous treatment effect in the equation (1) (i.e., *τ_L_* is a constant), but its overarching aim is to discern ‘individual’ treatment effect patterns. This is achieved by calculating a ‘forest weight’ for each sample, reflecting the frequency with which the other sample *q* co-occur in the same terminal node throughout the forest. Using the similar but weighted formats of the equation (1) by the forest weight, GRF estimates individual treatment effect of the target sample *p*, *τ_p_*, as follows:

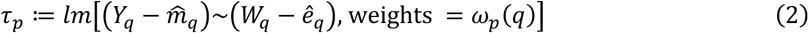

where *ω_p_*(*q*) denotes the forest weight vector of the target sample *p*. To prevent double-dipping calculation of *τ_p_*, GRF employs a unique algorithm called ‘honesty tree’^39, 42, 88, 89^, ensuring that only trees not using the sample *p* during tree construction engage in the calculation of *ω_p_*(*q*). This approach secures a rigorous computation of each sample’s individual treatment effect without any explicit data splitting.

Estimated individual treatment effects are instrumental in computing both the point estimate and standard error of the average treatment effect. GRF incorporates the Augmented Inverse Probability Weighting (AIPW) method, yielding doubly robust treatment effect estimates that is consistent even when either the outcome prediction model or the treatment model (i.e., *m* or *e*) is inaccurately specified. By adding a debiasing term to the estimated individual treatment effect, *τ*, GRF computes AIPW estimates of individual treatment effect, *τ^AIPW^*, with the following equation:

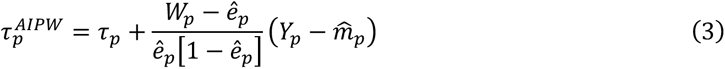

where *p* indicated a sample index. With data acquisition site specified as a clustering variable, the average treatment effect, *ATE*, is derived from cluster-level descriptive statistics of *τ^AIPW^* as follows:

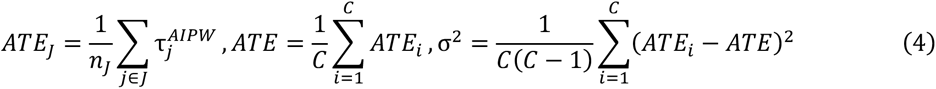

where *j* is a sample, *J* is a cluster, *n_J_* is the number of samples in the cluster *J*, and *C* is the total number of clusters, and σ^2^ is standard error of average treatment effect. Since GRF’s estimates follow Gaussian distribution in consistent and asymptotic manner^39, 89^, the point estimate and standard error of ATE also provides confidence interval for statistical testing.

#### Implementation of Generalized Random Forest

##### Model fitting and backward stepwise tuning

In this study, we constructed models to analyze both the acute and sustained effects of perceived discrimination on children’s stress levels, using outcome variables from timepoints CV4 and CV7, respectively. Given the substantial number of covariates relative to our sample size, we prioritized optimizing the models to ensure robust estimation of average and individual treatment effects since high dimensionality can lead to underpowered estimation^42^. Our model fitting commenced with the construction of GRF models incorporating all covariates under five different random seeds. Each model was built with 5,000 trees, and all adjustable hyperparameters (e.g., the minimum terminal node size) were fine-tuned using the ‘tune.parameters’ argument within the ‘causal_forest’ function. To mitigate potential site effects from the data collection sites, we used the ABCD study’s data acquisition sites as a clustering variable following the ref^41^. Subsequently, we aggregated the models from the five seeds into a ‘big forest’ model using the ‘merge_forest’ function. This aggregation aimed to enhance the robustness of our findings, ensuring that they were not overfitted to specific seeds. We then assessed the variable importance for each covariate through the ‘variable_importance’ function. The variable with the lowest importance was excluded, and these procedures were repeated until only one covariate was left. This iterative process resulted in 250 models for acute and sustained effects, respectively.

To determine the optimal model, we utilized a calibration test, which evaluated each model’s ability to accurately estimate average (*β_ATE_*) and individual treatment effects (*β_ITE_*). This test quantified these metrics with the following equation:

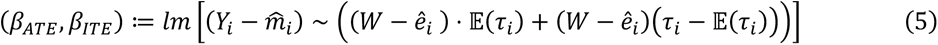

If *β_ATE_* and *β_ITE_* are close to 1, it means that average and individual treatment effect is well estimated even without the AIPW debiasing term^42^. A model passes this calibration test if it demonstrated significant *β_ATE_* and *β_ITE_* values (*P*_FDR_ < .05), as calculated by the ‘test_calibration’ function. From the models meeting this standard, the ‘best model’ was identified as the one with the lowest fit index, which reflects the sum of deviations of both calibration metrics from the ideal value of 1 as follows:

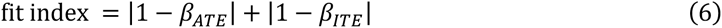

In the selection process, we adhered to the overlap assumption by excluding any model where predicted propensity scores were outside the .05 to .95 range.

### Estimation and aggregation of average treatment effect

We proceeded to estimate average treatment effect for each model. Using the ‘average_treatment_effect’ function, we calculated the point estimate of average treatment effect and its standard error per model. To synthesize these individual model estimates into a comprehensive assessment of average treatment effect, we employed a meta-analytic approach. Each model’s weight was computed as the inversed variance of average treatment effect estimates. Based on these weights, we combined the point estimates and standard errors from the 250 individual models using the following equation:

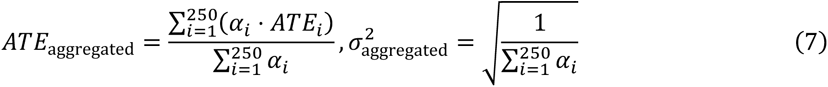

where *i* represents the model index and α signifies the weight assigned to each model’s estimate. The significance of the aggregated average treatment effect was then determined by verifying whether its 95% confidence interval excluded zero. Meta-analyses were performed for the 250 acute and 250 sustained models, respectively.

### Evaluation of individual variability in stress responses

We evaluated the existence of individual differences in acute and sustained effects of the perceived discrimination through twofold analyses. Firstly, we checked whether any of the 250 models each for acute or sustained effect analysis exhibited significant *β_ITE_*. This approach provides initial evidence of heterogeneity in individual treatment effects^42^. However, it should be noted that non-significant *β_ITE_* does not guarantee the absence of individual variations because non-significant *β_ITE_* would be also a signal of suboptimum. In other words, statistical testing of *β_ITE_* is a joint test of existence of heterogeneity and model optimization to detect heterogeneity. While 51 models for acute effects passed this examination, none of models for sustained effects did. Therefore, only with the best acute model, we performed the next analysis, called sorted group average treatment effect (GATE) test.

In the GATE test, we grouped samples based on quintiles of acute individual treatment effects calculated in the equation (2), using the ‘predict’ function. Then, we estimated group-level average treatment effects for each group through ‘average_treatment_effect’ function. We conducted one-tailed t-test (*P*_FDR_ < .05) to examine whether the any group’s average treatment effect was significantly higher than the estimates of the most resilient group (Q1). We followed the prior studies that assessed heterogeneity of the treatment effects based on GRF in the observational settings (e.g., ref^43, 44^).

### Identification of key vulnerability and resilience factors

We identified the covariates mainly engaging in acute stress vulnerability (high individual treatment effects) and resilience (low individual treatment effects) with the best acute model via triangulated analyses. Initially, we performed a group comparison test as a univariate analysis. This test examined statistical differences in 19 covariates incorporated in the acute best model between the most resilient (Q1) and the most vulnerable (Q5) groups. Statistical significance was assessed through a two-tailed t-test (*P*_FDR_ < .05). Next, we employed the best linear projection. This multiple linear regression analysis regressed acute individual treatment effects against 19 covariates. Using the ‘best_linear_projection’ function, we estimated the coefficients for each covariate within this model and evaluated their statistical significance (*P* < .05).

Based on the outcomes of both analyses, we categorized the covariates according to their type and tier. In terms of types, covariates were classified as either ‘resilience’ factors (significantly higher in Q1 than Q5 in the group comparison or displaying a significant negative coefficient in the best linear projection) or ‘vulnerability’ factors (significantly higher in Q5 than Q1 in the group comparison or showing significant positive coefficient in the best linear projection). Additionally, we assigned tiers to the covariates, with ‘tier 1’ representing significant results in both analyses, ‘tier 2’ indicating significant results in either the group comparison or the best linear projection, and ‘non-significant factor’ comprising non-significant results in both analyses.

Lastly, we conducted a partial dependence simulation to confirm the relationship between acute effects and individual covariates without assuming linearity of best linear projection while handling interactions among variables challenging to reflect in group comparison analysis. For each covariate, we generated 100 synthetic samples possessing all percentile values of the given variable while fixing the remaining 18 variables at their median values. Inputting these samples into the best acute model, we investigated the pattern of simulated acute individual treatment effects as the percentile value of the target covariate varies. All GRF analyses were implemented by the grf package^90^ (version 2.3.0) in the R studio (version 2023.03.1+446).

### Reproducibility assessment with the held-out dataset

#### The held-out samples

Among 11,879 samples in the ABCD dataset, we extracted 3,895 samples with demographic information and selected 19 neurostructural variables in the main analysis, treatment, and outcome variables. Afterward, we removed 1,116 samples used in the main GRF analysis, resulting in 2,779 samples. We randomly excluded one sibling within the identical biological family. Consequently, 2,503 children were determined as the final held-out dataset.

As in the main analysis, we imputed the missing values in the covariates with the k-nearest neighborhood approach and standardized the continuous variables. The treatment variable was binarized in the same way with the original dataset.

#### Replicating a detection of individual variability in acute stress response

The first aim of the reproducibility assessment was to examine whether we could detect individual variability in acute effects of perceived discrimination with the selected 19 covariates. For this, we constructed GRF models with 19 covariates of the held-out dataset with the backward stepwise tuning approach. Firstly, we examined whether any resulting models passed the calibration test (i.e., showing significant *β_ATE_* and *β_ITE_*) and chose the best acute model in terms of fit index (see equation (6)). Secondly, with the best model, we conducted a quintile-based GATE test. An identical approach was applied in the calibration and GATE tests with the main GRF analyses.

#### Replicating tiers and types of the frontotemporal factors

The second purpose of the reproducibility assessment was to investigate whether the selected 19 covariates displayed similar tiers and types in the held-out dataset with the original ones. We again characterized each covariate’s tiers and types through group comparison and best linear projection. We calculated Cramer’s *V* between results from two datasets, each for tier and type, to test their association between datasets. Due to the limited number of covariates, we evaluated the relationship between the two datasets only regarding the effect size of Cramer’s *V*, not its statistical significance.

### Sparse Generalized Canonical Correlation Analysis

We employed Sparse Generalized Canonical Correlation Analysis (sgCCA) to examine the multivariate relationship among each child’s neurostructural, environmental, and polygenic attributes. sgCCA is a dimension reduction technique that aims to maximize correlation coefficients among variable sets. The neurostructural set comprised 19 neural covariates, included in the best acute model. Concurrently, the environmental and polygenic sets consisted of variables measured at the same baseline time point, potentially interacting with the neural variables. The environmental set comprised nine variables, while the polygenic set comprised four.

To mitigate the potential confounding effects of demographic characteristics on neurostructural and environmental features, we regressed out the variance explained by factors such as sex, born overseas, race, age, acquisition site, parental income, and final degree for these variables. Polygenic scores were not subjected to this regression since demographic characteristics were not expected to influence genetic backgrounds. Subsequently, all residualized variables were standardized before the primary sgCCA model fitting.

We split the dataset into ‘discovery’ (*N* = 560) and ‘replication’ (*N* = 556) sets. We stratified both datasets regarding the quintile-based groups used in the GATE test so that each set includes the identical proportion of five groups as much as possible. With the discovery set, we explored the optimal sparsity hyperparameters through 1,000 times of permutation tests. In the permutation test, the squared sum of correlation coefficients among three blocks, *t*, was calculated for every sgCCA model. Given the null distribution of *t* obtained in the permutation test, the *P*-value of the original model, *P*_permutation_, was defined as follows:

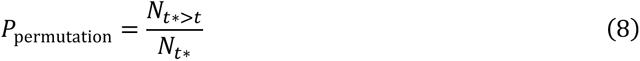

where *t*^∗^ and *t* are the squared sum of correlation coefficients from the permuted and original models, respectively, and *N* denotes the number of models. We chose the hyperparameter set that produced the lowest P-value in this test using the ‘rgcca_permutation’ function.

Using the best parameters selected in the discovery set, we fitted the sgCCA model again with the replication set. Firstly, we tested the statistical significance of the constructed model using the equation (8) through a 1,000 times permutation test. Secondly, we estimated the individual variable’s loading in each block and examined their statistical significance at *P*_FDR_ < .05. To obtain the confidence interval of each loading, we performed 1,000 times bootstrapping through the ‘rgcca_bootstrap’ function. All sgCCA modelings were conducted using the rgcca package^91^ (version 3.0.1) in the R studio (version 2023.03.1+446).

## Data Availability

All data produced are available online at

http://abcdstudy.org

## Acknowledgement

This work was supported by the National Research Foundation of Korea (NRF) grant funded by the Korea government (MSIT) (No. 2021R1C1C1006503, 2021K1A3A1A2103751212, 2021M3E5D2A01022515, RS-2023-00250759, RS-2023-00266787, RS-2023-00265406), by Creative-Pioneering Researchers Program through Seoul National University (No. 200- 20230058), by Semi-Supervised Learning Research Grant by SAMSUNG (No. A0426- 20220118), by Identify the network of brain preparation steps for concentration Research Grant by LooxidLabs (No. 339-20230001), by Institute of Information & communications Technology Planning & Evaluation (IITP) grant funded by the Korea government (MSIT) [NO.2021-0-01343, Artificial Intelligence Graduate School Program (Seoul National University)] and by the National Supercomputing Center with supercomputing resources including technical support (KSC-2022- CRE-0505).

## Supplementary Materials

**Supplementary Table 1.** Initially selected covariates (The neuroanatomical variables were not displayed for the legibility)

| Variable | Measurement | Time | Response | In GRF analysis? | In sgCCA modeling? |
| --- | --- | --- | --- | --- | --- |
| **Pre-COVID: demographic information** |  |  |  |  |  |
| Age | - | Baseline | Parent | O | X |
| Born overseas | - | Baseline | Parent | O | X |
| Race | - | Baseline | Parent | O | X |
| Sex | - | Baseline | Parent | O | X |
| Parental income | - | Baseline | Parent | O | X |
| Parental final degree | - | Baseline | Parent | O | X |
| **Pre-COVID: psychological features** |  |  |  |  |  |
| Internalizing problem syndrome score | Child Behavior Checklist – ‘CBCL’<sup>1</sup> | 3-year-follow | Parent | O | X |
| Externalizing problem syndrome score | CBCL<sup>1</sup> | 3-year-follow | Parent | O | X |
| Total problem syndrome score | CBCL<sup>1</sup> | 3-year-follow | Parent | O | X |
| Depressive disorder symptom score | CBCL<sup>1</sup> | 3-year-follow | Parent | O | X |
| Anxiety disorder symptom score | CBCL<sup>1</sup> | 3-year-follow | Parent | O | X |
| Stress score | CBCL<sup>1</sup> | 3-year-follow | Parent | O | X |
| Emotion regulation strategy: not showing | Emotion Regulation Questionnaire – ‘ERQ’<sup>2</sup> | 3-year-follow | Child | O | X |
| Emotion regulation strategy: change the way | ERQ<sup>2</sup> | 3-year-follow | Child | O | X |
| Emotion regulation strategy: change the object | ERQ<sup>2</sup> | 3-year-follow | Child | O | X |
| **Pre-COVID: environmental features** |  |  |  |  |  |
| Parental monitoring score | ref<sup>3, 4</sup> | Baseline | Child | O | O |
| Family conflict score | Family Environment Scale<sup>5</sup> | Baseline | Child | O | O |
| Prosocial behavior score | Strengths and Difficulties Questionnaire<sup>6</sup> | Baseline | Child | O | O |
| Parental acceptance score | Child Report of Behavior Inventory<sup>7, 8</sup> | Baseline | Child | O | O |
| Neighborhood safety rating | ref<sup>9, 10</sup> | Baseline | Parent | O | O |
| Traumatic event: car accident | Kiddie Schedule for Affective Disorders and Schizophrenia – ‘KSADS’<sup>11</sup> | Baseline | Parent | X<br>(Near zero variance) | X |
| Traumatic event: another accident | KSADS<sup>11</sup> | Baseline | Parent | X<br>(Near zero variance) | X |
| Traumatic event: caught in a fire | KSADS<sup>11</sup> | Baseline | Parent | X<br>(Near zero variance) | X |
| Traumatic event: caught in a natural disaster | KSADS<sup>11</sup> | Baseline | Parent | X<br>(Near zero variance) | X |
| Traumatic event: terrorism | KSADS<sup>11</sup> | Baseline | Parent | X<br>(Near zero variance) | X |
| Traumatic event: mass destruction | KSADS<sup>11</sup> | Baseline | Parent | X<br>(Near zero variance) | X |
| Traumatic event: witnessing a shot/stabbing | KSADS<sup>11</sup> | Baseline | Parent | X<br>(Near zero variance) | X |
| Traumatic event: shot/stabbed/beaten by a non-family member | KSADS<sup>11</sup> | Baseline | Parent | X<br>(Near zero variance) | X |
| Traumatic event: shot/stabbed/beaten by a family member | KSADS<sup>11</sup> | Baseline | Parent | X<br>(Near zero variance) | X |
| Traumatic event: having bruises by a family member | KSADS<sup>11</sup> | Baseline | Parent | X<br>(Near zero variance) | X |
| Traumatic event: threatened to kill you by a non-family member | KSADS<sup>11</sup> | Baseline | Parent | X<br>(Near zero variance) | X |
| Traumatic event: threatened to kill you by a family member | KSADS<sup>11</sup> | Baseline | Parent | X<br>(Near zero variance) | X |
| Traumatic event: witnessing the grown-ups in the home push, shove or hit one another | KSADS<sup>11</sup> | Baseline | Parent | X<br>(Near zero variance) | X |
| Traumatic event: sexually harassed by a family member | KSADS<sup>11</sup> | Baseline | Parent | X<br>(Near zero variance) | X |
| Traumatic event: sexually harassed by a non-family member | KSADS<sup>11</sup> | Baseline | Parent | X<br>(Near zero variance) | X |
| Traumatic event: sexually harassed by a peer | KSADS<sup>11</sup> | Baseline | Parent | X<br>(Near zero variance) | X |
| Traumatic event: learned about the sudden death of a loved one | KSADS<sup>11</sup> | Baseline | Parent | O | O |
| Discrimination: not feel accepted by other Americans | Measure of Perceived Discrimination<sup>12</sup> | 2-year-follow | Child | X<br>(Near zero variance) | X |
| Racial identity: important to develop the practices of my heritage culture | Vancouver Index of Acculturation – ‘VIA’<sup>13</sup> | 3-year-follow | Child | X<br>(too many NAs) | X |
| Racial identity: important to develop American mainstream cultural practices | VIA<sup>13</sup> | 3-year-follow | Child | X<br>(too many NAs) | X |
| **Pre-COVID: multiple ancestries-based polygenic scores** |  |  |  |  |  |
| Depression | Supplementary Note 1 | Baseline | Child | O | O |
| Posttraumatic stress disorder | Supplementary Note 1 | Baseline | Child | O | O |
| Schizophrenia | Supplementary Note 1 | Baseline | Child | O | O |
| Body mass index | Supplementary Note 1 | Baseline | Child | O | O |
| **Post-COVID: environmental & socioeconomic features** |  |  |  |  |  |
| Participation in the family activities | ABCD COVID Rapid Response Research Survey – ‘RRR’ | CV2 | Child | X<br>(Near zero variance) | X |
| Relationships with the family | COVID RRR | CV2 | Child | X<br>(Near zero variance) | X |
| Perceived economic difficulties during the pandemic | COVID RRR | CV2 | Parent | O | X |
| Wage loss during the pandemic | COVID RRR | CV2 | Parent | O | X |
| Child’s history of COVID-19 diagnosis | COVID RRR | CV2 | Parent | X<br>(Near zero variance) | X |
| County-level cumulative COVID-19 cases | COVID RRR | CV2 | Geocoded | O | X |
| County-level cumulative COVID-19 deaths | COVID RRR | CV2 | Geocoded | O | X |
| County-level unemployment rate | COVID RRR | CV2 | Geocoded | O | X |
| Median distance traveled from home during the pandemic | COVID RRR | CV2 | Geocoded | O | X |
| The average time at home during the pandemic | COVID RRR | CV2 | Geocoded | O | X |
| The number of electronic devices at home | COVID RRR | CV2 | Geocoded | O | X |
| Full-time working behavior | COVID RRR | CV2 | Geocoded | O | X |
| Part-time working behavior | COVID RRR | CV2 | Geocoded | O | X |

**Supplementary Table 2.**
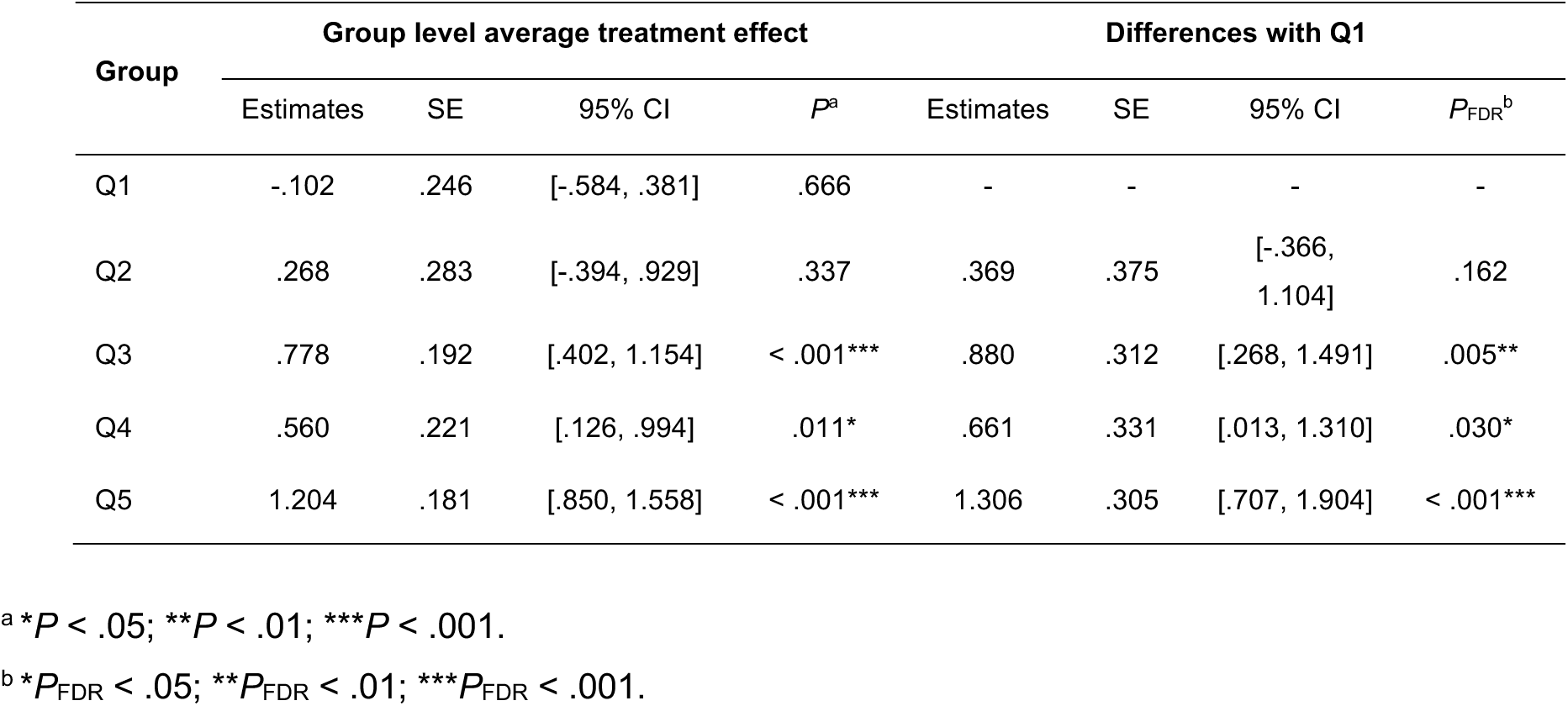
The results of GATE test as an assessment of individual variability in acute individual treatment effects.

**Supplementary Table 3.** The tiers and types of individual covariates selected in the best acute model based on the results of group comparison and best linear projection.

| Covariate | Group comparison |  |  |  | Best linear projection |  |  | Type | Tier |
| --- | --- | --- | --- | --- | --- | --- | --- | --- | --- |
| | Q1<sup>a</sup><br>Mean (SD) | Q5<sup>a</sup><br>Mean (SD) | *t* | *P*<sub>FDR</sub><sup>b</sup> | $\beta$ | SE | *P*<sup>c</sup> | | |
| VOL L PTR | .225 (1.2) | -.211 (.8) | 4.403 | < .001*** | -.325 | .107 | .003** | Resilience | 1 |
| SC L POP-L POR | .281 (1.4) | -.204 (.8) | 4.483 | < .001*** | -.311 | .135 | .022* | Resilience | 1 |
| SC L PHIG-L HI | .296 (1.5) | -.104 (.8) | 3.622 | < .001*** | -.357 | .132 | .007** | Resilience | 1 |
| VOL L PHIG | .101 (.9) | -.185 (1.2) | 2.820 | .007** | .044 | .113 | .697 | Resilience | 2 |
| VOL R PHIG | .176 (.9) | -.312 (1.2) | 4.844 | < .001*** | -.193 | .135 | .155 | Resilience | 2 |
| SC L CMFG-R POP | .400 (1.4) | -.185 (.8) | 5.315 | < .001*** | -.183 | .217 | .400 | Resilience | 2 |
| SC L MOFG-R POR | .316 (1.5) | -.301 (.6) | 5.792 | < .001*** | -.183 | .245 | .456 | Resilience | 2 |
| SC L MOFG-R RMFG | .156 (1.0) | -.314 (.9) | 5.282 | < .001*** | -.167 | .123 | .177 | Resilience | 2 |
| SC L POR-L PTR | .042 (1.1) | -.209 (.9) | 2.608 | .012* | .194 | .156 | .214 | Resilience | 2 |
| SC L POR-L RMFG | .255 (1.1) | -.436 (.9) | 7.043 | < .001*** | -.274 | .162 | .092 | Resilience | 2 |
| SC L RMFG-R LOFG | .172 (1.3) | -.220 (.9) | 3.694 | < .001*** | -.252 | .239 | .292 | Resilience | 2 |
| SC R POR-R RMFG | .391 (1.2) | -.505 (.8) | 9.059 | < .001*** | .036 | .166 | .827 | Resilience | 2 |
| SC L MOFG-L PTR | -.180 (.9) | .320 (1.2) | -5.026 | < .001*** | .189 | .249 | .447 | Vulnerability | 2 |
| SC L PTR-L SFG | -.307 (1.0) | .250 (1.0) | -5.711 | < .001*** | .070 | .108 | .517 | Vulnerability | 2 |
| SC R MOFG-R PTR | -.292 (.6) | .560 (1.6) | -7.383 | < .001*** | .179 | .121 | .140 | Vulnerability | 2 |
| VOL R HI | .053 (1.1) | -.09 (.9) | 1.514 | .155 | .252 | .162 | .120 | n.s. | n.s. |
| SC L CMFG-R SFG | .051 (1.0) | -.004 (1.1) | .555 | .579 | .089 | .148 | .549 | n.s. | n.s. |
| SC L POP-L RMFG | -.067 (1.1) | .035 (.9) | -1.046 | .313 | .025 | .138 | .859 | n.s. | n.s. |
| SC R EG-R PHIG | .100 (1.0) | -.010 (1.0) | 1.168 | .272 | .027 | .160 | .865 | n.s. | n.s. |

<sup>a</sup> All values were standardized.<sup>b</sup> \**P*<sub>FDR</sub> < .05; \*\**P*<sub>FDR</sub> < .01; \*\*\**P*<sub>FDR</sub> < .001.<sup>c</sup> \**P* < .05; \*\**P* < .01; \*\*\**P* < .001.

**Supplementary Table 4.**
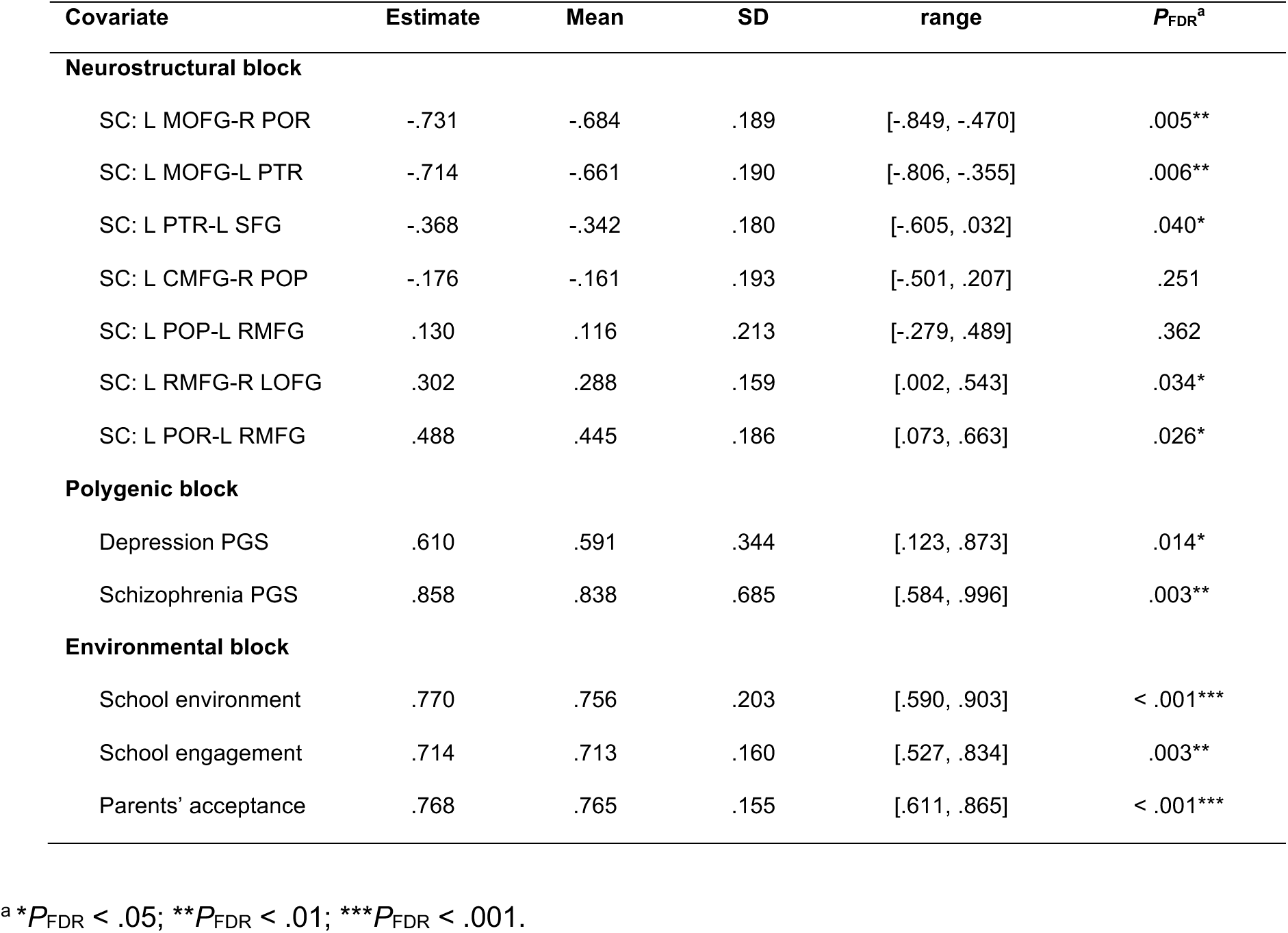
The loadings of individual variables in the sgCCA model obtained from 1,000 times bootstrapping.

| Covariate | Estimate | Mean | SD | range | $P_{FDR}^a$ |
| --- | --- | --- | --- | --- | --- |
| **Neurostructural block** |  |  |  |  |  |
| SC: L MOFG-R POR | -.731 | -.684 | .189 | [-.849, -.470] | .005** |
| SC: L MOFG-L PTR | -.714 | -.661 | .190 | [-.806, -.355] | .006** |
| SC: L PTR-L SFG | -.368 | -.342 | .180 | [-.605, .032] | .040* |
| SC: L CMFG-R POP | -.176 | -.161 | .193 | [-.501, .207] | .251 |
| SC: L POP-L RMFG | .130 | .116 | .213 | [-.279, .489] | .362 |
| SC: L RMFG-R LOFG | .302 | .288 | .159 | [.002, .543] | .034* |
| SC: L POR-L RMFG | .488 | .445 | .186 | [.073, .663] | .026* |
| **Polygenic block** |  |  |  |  |  |
| Depression PGS | .610 | .591 | .344 | [.123, .873] | .014* |
| Schizophrenia PGS | .858 | .838 | .685 | [.584, .996] | .003** |
| **Environmental block** |  |  |  |  |  |
| School environment | .770 | .756 | .203 | [.590, .903] | < .001*** |
| School engagement | .714 | .713 | .160 | [.527, .834] | .003** |
| Parents' acceptance | .768 | .765 | .155 | [.611, .865] | < .001*** |

<sup>a</sup> \* $P_{FDR} < .05$ ; \*\* $P_{FDR} < .01$ ; \*\*\* $P_{FDR} < .001$ .

**Supplementary Fig 1.**
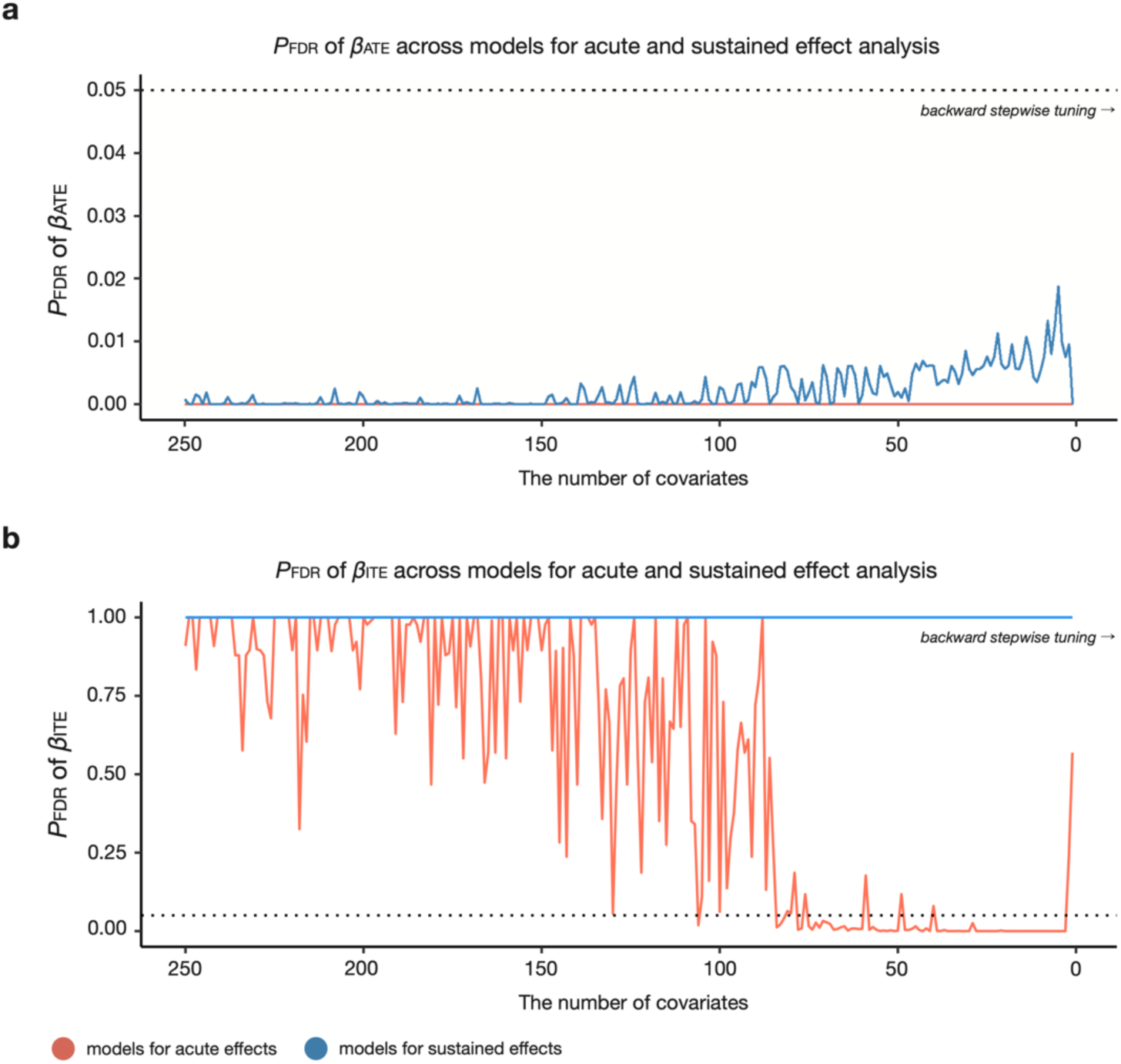
Statistical significance of calibration coefficients *β_ATE_* and *β*_ITE_ across GRF models. **a,** *P*_FDR_ of *β_ATE_*_’_ across GRF models for acute and sustained effect analysis. The dotted horizontal line denotes the criteria of statistical significance (*P*_FDR_ < .05). **b,** *P*_FDR_ of *β_ITE_* across models for acute and sustained effect analysis.

**Supplementary Fig 2.**
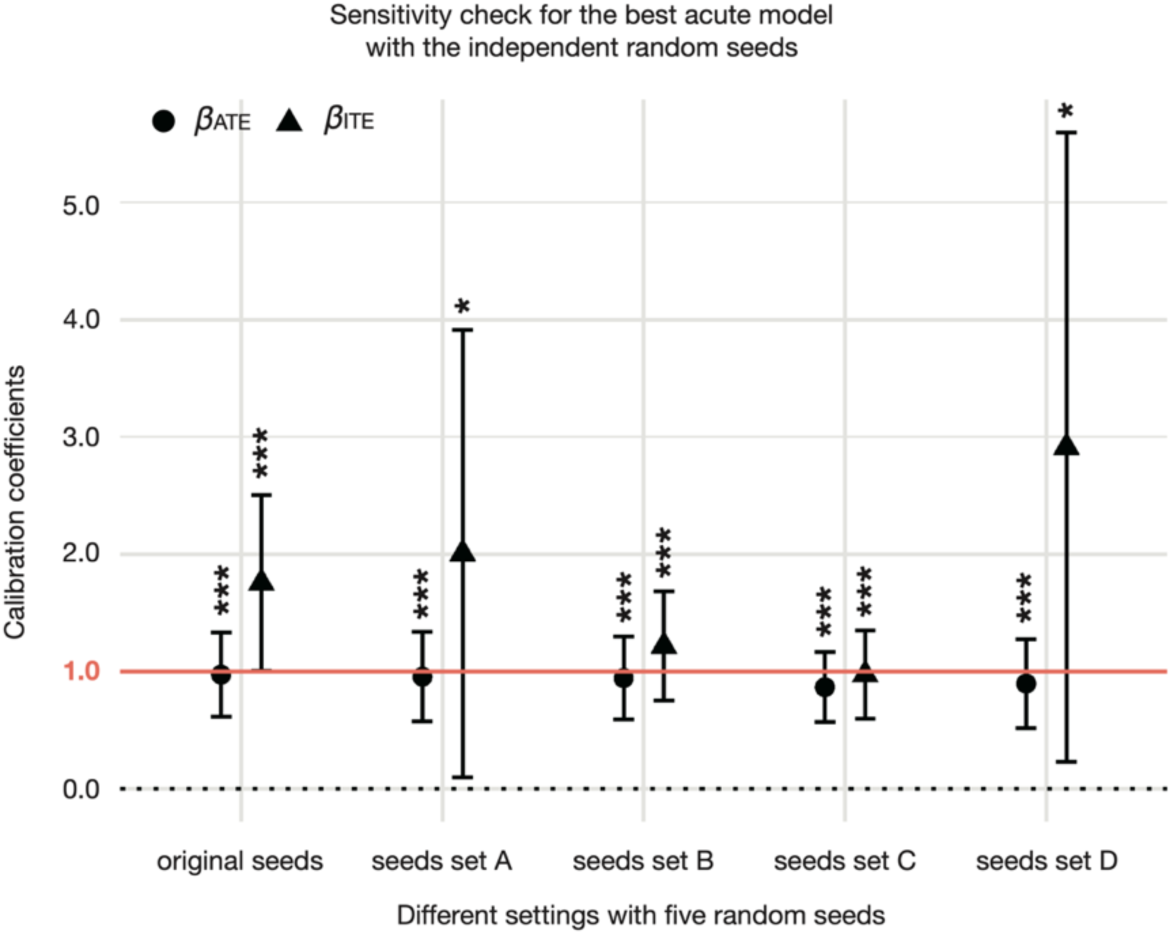
Replicating results of the best acute model in calibration test under independent random seeds. We examined whether the combination of 19 covariates chosen by the best acute model could pass the calibration test in the other random seed sets. This replication analysis aimed to test the possibility of overfitting the specific seed settings in the best acute model, although we aimed to prevent it via the seed ensemble approach. As we did in the main analysis, we generated four independent seed sets (i.e., seeds set A, B, C, and D), each consisting of five random seeds. We constructed the ‘big forest’ model with 19 covariates in each seed set and evaluated its model fit through a calibration test. Specifically, the significance of two model fit metrics (i.e., *β_ATE_* and *β_ITE_*) from the calibration test was assessed. The error bar denotes the 95% confidence interval. The dotted line indicates the null hypothesis of statistical testing, and the red solid line means the ideal value in the calibration test. Selected 19 covariates in the main GRF analysis produced the model with a modest performance to estimate average and individual treatment effects in all seed settings. \**P* < .05; \*\**P* < .01; \*\*\**P* < .001.

**Supplementary Fig 3.**
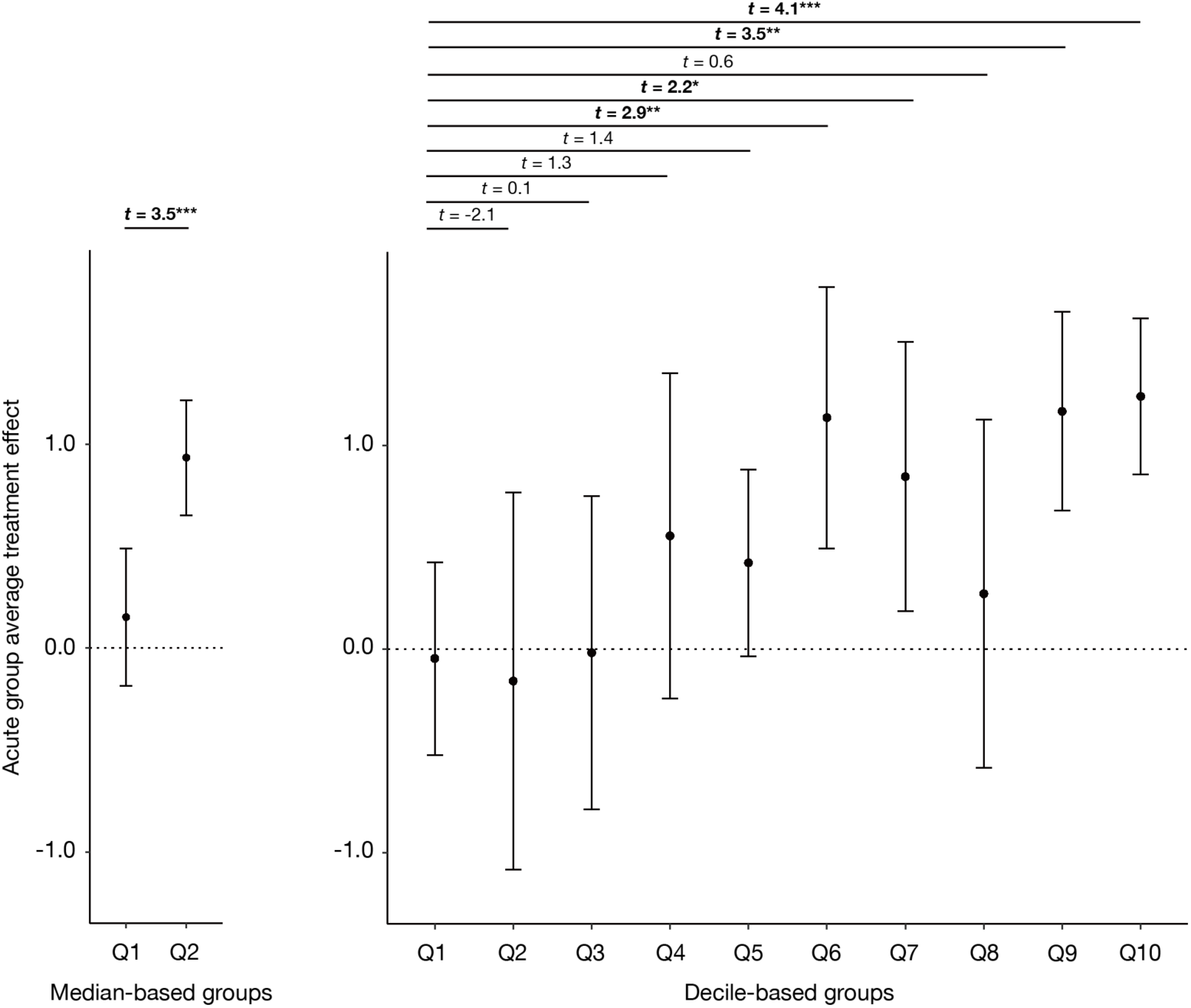
The results of the GATE test with the smaller and larger number of groups. We tested the variability of the acute treatment effects through the GATE test with a smaller and larger number of groups than quintile-based groupings. This analysis examined whether the observed polarized pattern in acute effects relies on the number of groups, the main parameter in the GATE test. The left and right panels denote the results of GATE tests based on two and ten groups, respectively. Relatively long confidence intervals and non-monotonic estimates of group-level average treatment effects in the decile-based GATE test may reflect the small sample size in each group (about 100 samples). Nevertheless, both results present the heterogeneous acute effects across groups, indicating the individual differences in acute stress response to perceived discrimination. The bolded statistics denote the statistically significant differences between groups. \**P*_FDR_ < .05; \*\**P*_FDR_ < .01; \*\*\**P*_FDR_ < .001.

**Supplementary Fig 4.**
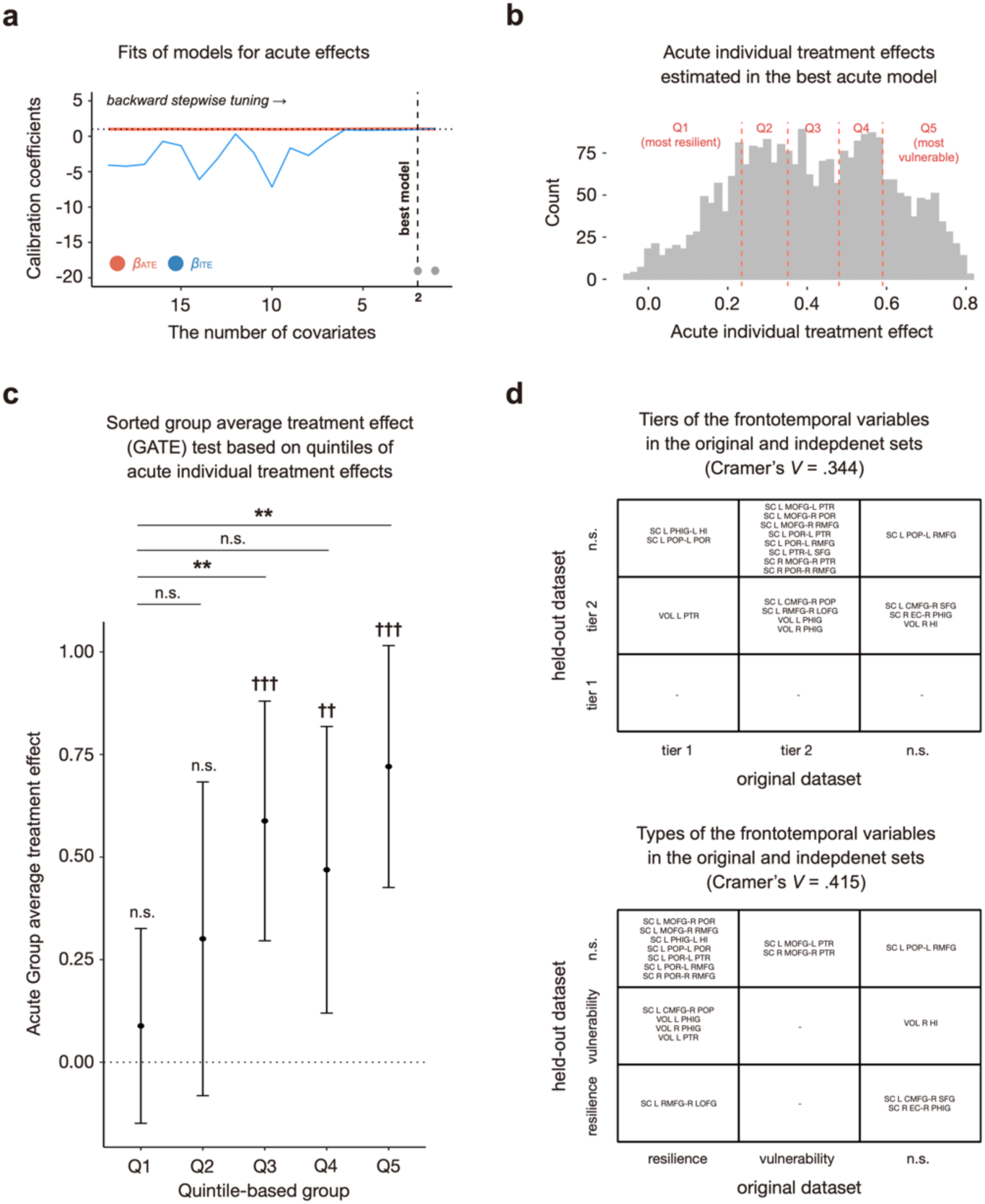
Reproducibility tests with the held-out dataset (*N* = 2,503) **a,** The model fit evaluation through calibration test with the held-out samples. We conducted backward stepwise tuning for GRF models with the 19 key neurostructural factors identified in the main analysis. From the resulting 19 models, we observed that the last two models accurately estimated both average and individual treatment effects of acute effects. **b,** The distribution of acute individual treatment effects estimated by the best model in the held-out samples. **c,** Results of GATE test to assess individual variability in acute individual treatment effects. The error bar denotes a 95% confidence interval of the group-level average treatment effect. Cross symbols above each error bar show the significance of group-level average treatment effects (☨*P* < .05; ☨☨*P* < .01; ☨☨☨*P* < .001). Asterisks above the bar plot indicate the significance of differences in group-level average treatment effects (\**P*_FDR_ < .05; \*\**P*_FDR_ < .01; \*\*\**P*_FDR_ < .001). **d,** The tiers and types of each neurostructural covariate in the original and held-out dataset.

**Supplementary Fig 5.**
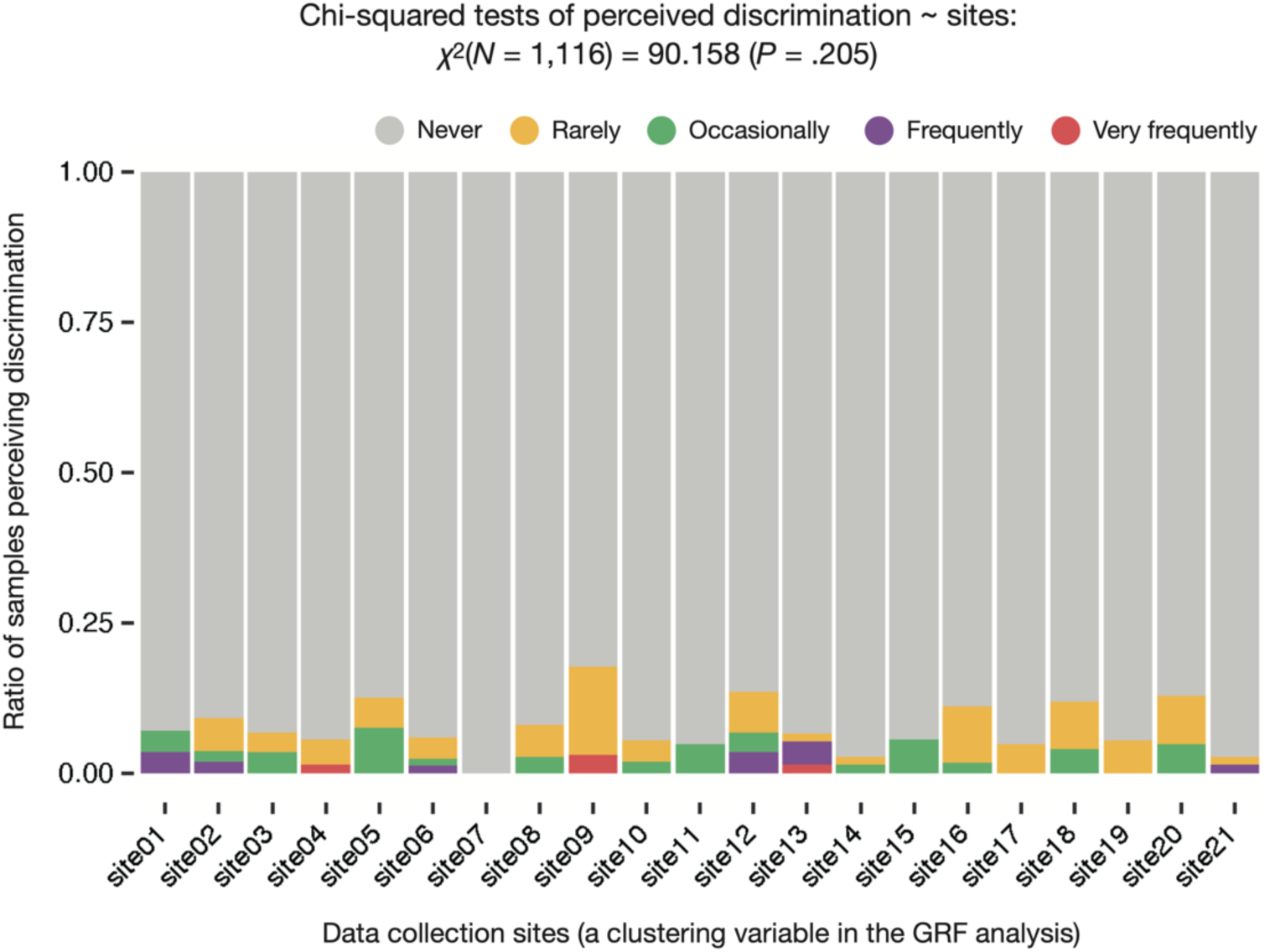
Perceived COVID-19-related discrimination across data collection sites. We performed a chi-squared test to examine the statistical differences in the frequency of perceiving discrimination across data collection sites. No significant difference in the frequency was detected in our dataset (χ^2^ = 90.158, *P* = .253).

## Supplementary Note 1. Genotyping, computation, and validation of multiple ancestries-based polygenic scores

The ABCD study genotyped participants’ saliva samples using Rutgers University Cell and DNA Repository, including 733,293 single nucleotide polymorphisms (SNPs). These SNPs were filtered through standard PLINK criteria as follows: genotype call rate < 95% removed, sample call rate < 95% removed, and minor allele frequency < 1% removed. Afterward, we imputed genotypes with Eagle v2.4^14^ and the Michigan Imputation Sever^15^. For robust quality control, we again removed SNPs with the following conditions: INFO score < 0.4 removed, genotype call rate < 95% removed, Hardy-Weinberg Equilibrium *p*-value < 1×10^-20^ removed, sample missingness > 5% removed, minor allele frequency < 0.5% removed, and extreme heterozygosity (i.e., over three standard deviations of the population mean). As a result, a total of 11,221,810 SNPs variants remained.

Considering that the ABCD study dataset includes samples with diverse ethnic backgrounds, we relaxed possible population bias from genetic relatedness and ancestry mixture. To identify genetically unrelated individuals, we estimated kinship coefficients and principal components of ancestral information using PC-Air^16^ and PC-Relate^17^. We labeled a sample as ‘unrelated individuals’ if he or she satisfied the following criteria: kinship coefficients > 0.022 and over six standard deviations of the population mean in the principal component space. Consequently, final genotyped samples of 8,620 unrelated children were included in the main analysis, and an independent set of 1,579 individuals was used for validating polygenic scores.

We calculated children’s polygenic scores for four distinct traits associated with stress vulnerability with GWAS summary statistics based on discovery samples from multiple ancestries: post-traumatic stress disorder^18^ (PTSD), schizophrenia^19, 20^ (SCZ), depression^21, 22^ (DEP), and body mass index^23, 24^ (BMI). With these GWAS summary statistics as input, we used PRS-CSx^25^, a high-dimensional Bayesian regression approach utilizing shrinkage prior to estimating the posterior effect sizes of SNPs on the complex traits. This approach is more specialized in incorporating multiethnic populations and showed higher predictive power in simulation and empirical data analysis than the other algorithms^25^.

In the held-out set of 1,579 unrelated children, validation for hyperparameter optimization was conducted. These unrelated samples consist of 88 of African ancestry, 25 of East Asian ancestry, 1,365 of European ancestry, 88 of American ancestry, and 55 not specified. We manually explored the global shrinkage parameter that maximizes the explanatory power of the regression model and effect sizes of polygenic scores on the target phenotype. Following the recommendation of the ref.^25^, we conducted a grid search to optimize a shrinkage parameter by testing 1×10^-6^, 1×10^-4^, 1×10^-2^, and 1 within the validation set. We built single ancestry-based polygenic scores and multiple ancestries-based polygenic scores for each trait. For example, if GWAS from two populations were available, three polygenic scores could be generated, two from each population and one by incorporating both populations. Each model regressed the related phenotype from the ABCD study on the polygenic score(s), sex, top 10 principal components of genotype data, and genetic ancestry identified by ADMIXTURE algorithm^26^. This regression-based validation was conducted only with polygenic scores of PTSD, DEP, and BMI that had corresponding measures in the ABCD study dataset. In this model, we defined multiethnic polygenic scores as a linear combination of two polygenic predictors (i.e., European-based and ancestry-specific polygenic scores). Finally, we chose the best shrinkage parameters (either multiple ancestries-based polygenic scores or combinations of single ancestry-based polygenic scores) for each trait based on the R-squared of the regression model and the effect sizes (beta coefficient) of polygenic scores. The other polygenic scores, ALCDEP and SCZ, were automatically validated by PRS-CS-auto^25^, in which the optimal shrinkage parameter was chosen through a fully Bayesian approach. All polygenic scores were adjusted for sex, age, study site, and genetic ancestry.

## Supplementary Note 2. Estimating streamline counts from diffusion-weighted imaging data

We used diffusion-weighted imaging (DWI) data measured at the baseline timepoint in the ABCD study dataset. DWI data were preprocessed by the ABCD Data Analysis and Informatics Center following the ABCD-specific protocols^27^. In brief, Eddy current distortion correction was performed to predict the overall pattern of artifact distortions^28^. Corrected images were registered to images synthesized from tensor fit^[29]^ and adjusted with diffusion gradients^27, 30^ to minimize noises from head motions. Then, diffusion tensors were estimated to identify and remove slices distorted by abrupt head motion^31^. Using the ‘TOPUP’ function in FSL^32, 33^, B0 distortions were corrected via the reversing gradient method. After gradient nonlinearity distortion correction^34^, images of b=0 were registered T1w-images based on their mutual information^35^. Finally, corrected images were resampled into a standard orientation with 1.7mm isotropic resolution.

To estimate streamline counts among ROIs – potential neural sources of stress resilience and vulnerability, we first estimated each child’s structural connectome. We applied ‘MRtrix3^36^’to generate individual probabilistic tractography. Then, we estimated streamline counts from the resulted tractograms following the ref^37–40^. We estimated the noise maps and computed the objective threshold on the eigenvalues to conduct principal component analysis (PCA) for the noise level-based denoising^41^. We also performed bias correction using ANTs’ N4 algorithm^42^. Afterward, we obtained probabilistic tractography by second-order integration over fiber orientation distributions with random seeds across the brain and streamline counts of 20 million^43^. These initial tractograms were filtered out preliminary tractograms with spherical-deconvolution filtering with 2:1 ratio. By doing so, we extracted 10 million streamline counts and 84×84 whole-brain connectome matrix with T1-based parcellation and segmentation. We performed these procedures with the supercomputers at Argonne Leadership Computing Facility Theta and Texas Advanced Computing Center Stampede2.

